# Vision-language framework for multi-sequence brain magnetic resonance imaging

**DOI:** 10.64898/2026.03.30.26349106

**Authors:** Diala Lteif, Shuyue Jia, Subhrangshu Bit, Artem Kaliaev, Asim Z. Mian, Juan E. Small, Balamurugan Mangaleswaran, Bryan A. Plummer, Sarah A. Bargal, Rhoda Au, Vijaya B. Kolachalama

**Affiliations:** Department of Computer Science, Boston University, MA, USA; Department of Electrical & Computer Engineering, Boston University, MA, USA; Department of Radiology, Boston University Chobanian & Avedisian School of Medicine, Boston, MA, USA; Department of Radiology, Lahey Hospital & Medical Center, Burlington, MA, USA; Brain and Spine Hospital, Chennai, India; Department of Computer Science, Georgetown University, Washington, DC, USA; Department of Medicine, Boston University Chobanian & Avedisian School of Medicine, Boston, MA, USA; Department of Neurology, Boston University Chobanian & Avedisian School of Medicine, Boston, MA, USA; Boston University Alzheimer’s Disease Research Center, Boston, MA, USA; The Framingham Heart Study, Boston University Chobanian & Avedisian School of Medicine, Boston, MA, USA; Department of Anatomy and Neurobiology, Boston University Chobanian & Avedisian School of Medicine, Boston, MA, USA; Department of Epidemiology, Boston University School of Public Health, Boston, MA, USA; Faculty of Computing & Data Sciences, Boston University, MA, USA

**Author notes:** Corresponding author: Vijaya B. Kolachalama, PhD.

## Abstract

Structural magnetic resonance imaging (MRI) is a cornerstone for diagnosing neurological disorders, yet automated interpretation of multi-sequence brain MRI remains limited by challenges in cross-sequence reasoning and protocol variability. Here we present ReMIND, a vision-language modeling framework tailored for comprehensive multi-sequence and multi-volumetric brain MRI analysis. Trained on over 73,000 deidentified patient visits encompassing more than 850,000 MRI sequences paired with radiology reports from diverse clinical and research cohorts, ReMIND combined large-scale instruction tuning on more than one million clinically grounded question–answer (QA) pairs with targeted supervised fine-tuning for radiology report generation. At inference, ReMIND employed modality-aware reranking and correction, a report-level decoding strategy that suppressed unsupported modality claims while preserving linguistic fluency and clinical coherence. Cross-cohort generalization was maintained on independent external datasets from different institutions. These findings represent an advance toward consistent and equitable brain MRI interpretation, meriting prospective evaluation to support diagnosis and management of neurological conditions.

## Introduction

Magnetic resonance imaging (MRI) plays an important role in the evaluation of a range of neurological disorders, including neurodegenerative, vascular, inflammatory, neoplastic and demyelinating conditions, particularly those with characteristic or detectable changes on imaging ^1–5^. In neurology, MRI enables systematic “ruling in” or “ruling out” of disease by revealing subtle morphologic changes, such as regional atrophy, white matter signal abnormalities, lesions or vascular anomalies, which serve as markers of underlying pathology. However, the practical and diagnostic impact of MRI is contingent on expert interpretation. The process of extracting clinically actionable meaning from MRIs requires more than just visual inspection; radiologists methodically analyze multi-sequence images, assess for focal and diffuse patterns, and evaluate findings against the clinical backdrop of patient history and presentation. This synthesis culminates in a structured report, where subtle imaging observations are integrated with diagnostic reasoning, yielding impressions that inform patient management. Such interpretive expertise is a product of extensive training and experience, and remains concentrated among a limited pool of highly specialized radiologists, whose availability is increasingly strained as the prevalence of neurological disorders rises ^6,7^. This scarcity contributes to disparities in timely and accurate diagnosis, and forms a significant barrier to the widespread and equitable use of advanced neuroimaging. To bridge the gap between MRI’s technological potential and clinical realities, scalable, standardized approaches are needed that can democratize expert-level assessment.

Artificial intelligence (AI) offers a promising avenue to address these challenges by augmenting the interpretation of MRI scans and streamlining diagnostic workflows. Specifically, AI-driven systems can be designed to assist radiologists or other providers by automating the detection of key imaging findings, while also generating structured, clinically meaningful reports ^8,9^. Unlike traditional AI models that provide raw image or single-slice analysis, advanced systems based on vision-language models (VLMs) combine imaging insights with contextual patient data, enabling outputs that mimic the integrative reasoning of expert radiologists. Such tools not only hold potential to reduce cognitive burden and improve diagnostic efficiency, but also to assist less specialized clinicians in managing complex neurological cases. Despite their promise, VLMs face limitations in brain MRI applications, such as handling multi-sequence data and diverse neurological conditions, due to data scarcity and task complexity. Recent progress in the development of foundation models and self-supervised vision-language learning has nonetheless accelerated possibilities in medical imaging ^10,11^. These multimodal models, pre-trained on large-scale, unannotated datasets and capable of jointly modeling images and associated free text, have achieved impressive results in tasks such as classification, retrieval, visual question-answering (VQA), segmentation, and automated radiology report generation (RRG) ^12–17^. Yet, development and validation of VLMs for multi-sequence brain MRI targeting various neurological conditions remain limited.

To address this gap, we introduce ReMIND (Radiology–encoded Multimodal Interpretation for Neurological Disorders), a vision-language modeling framework on a large-scale dataset of multi-sequence brain MRIs paired with diverse question-answer annotations and radiologist reports. Leveraging state-of-the-art foundation model architectures, ReMIND learned joint representations of imaging and language to generate structured, radiology-style reports aligned with standard clinical workflows. Also, we introduced an inference-time decoding strategy that operates at the report level to suppress unsupported modality claims while preserving linguistic fluency and clinical coherence. We rigorously evaluated ReMIND across clinical and research cohorts. The model demonstrated strong, generalizable performance across a range of neurological disorders. Together, these findings highlight the potential of VLMs for brain MRI analysis and set the stage for more accessible, consistent and structurally-informed diagnosis of neurological conditions.

## Results

The objective of this study was to develop and validate a vision-language model tailored for comprehensive interpretation of multi-sequence brain MRI in neurological disorders (Fig. 1). Drawing from thousands of de-identified patient visits encompassing various imaging sequences paired with radiology reports from diverse clinical and research cohorts (Table 1), ReMIND was trained through large-scale instruction tuning on clinically grounded question–answer pairs, followed by targeted supervised fine-tuning and a modality-aware faithful decoding mechanism designed to prevent unsupported modality claims in incomplete-protocol cases. We conducted comprehensive evaluations across visual question-answering (VQA) and radiology report generation (RRG) tasks to assess ReMIND’s domain adaptation, generalization and clinical utility (Fig. 2 & Table 2). Performance was benchmarked on internal held-out testing data from GH as well as external clinical and research cohorts, including Boston Medical Center, Boston, MA, USA (BMC), Lahey Hospital & Medical Center, Burlington, MA, USA (LHMC), Brain and Spine Hospital, Chennai, India (BSH), and National Alzheimer’s Coordinating Center (NACC). Evaluation employed a combination of standard classification and language-level metrics, with particular emphasis on reducing modality hallucination rates under natural and controlled ablation conditions through a novel decoding strategy (Fig. 3).

**Figure 1:**
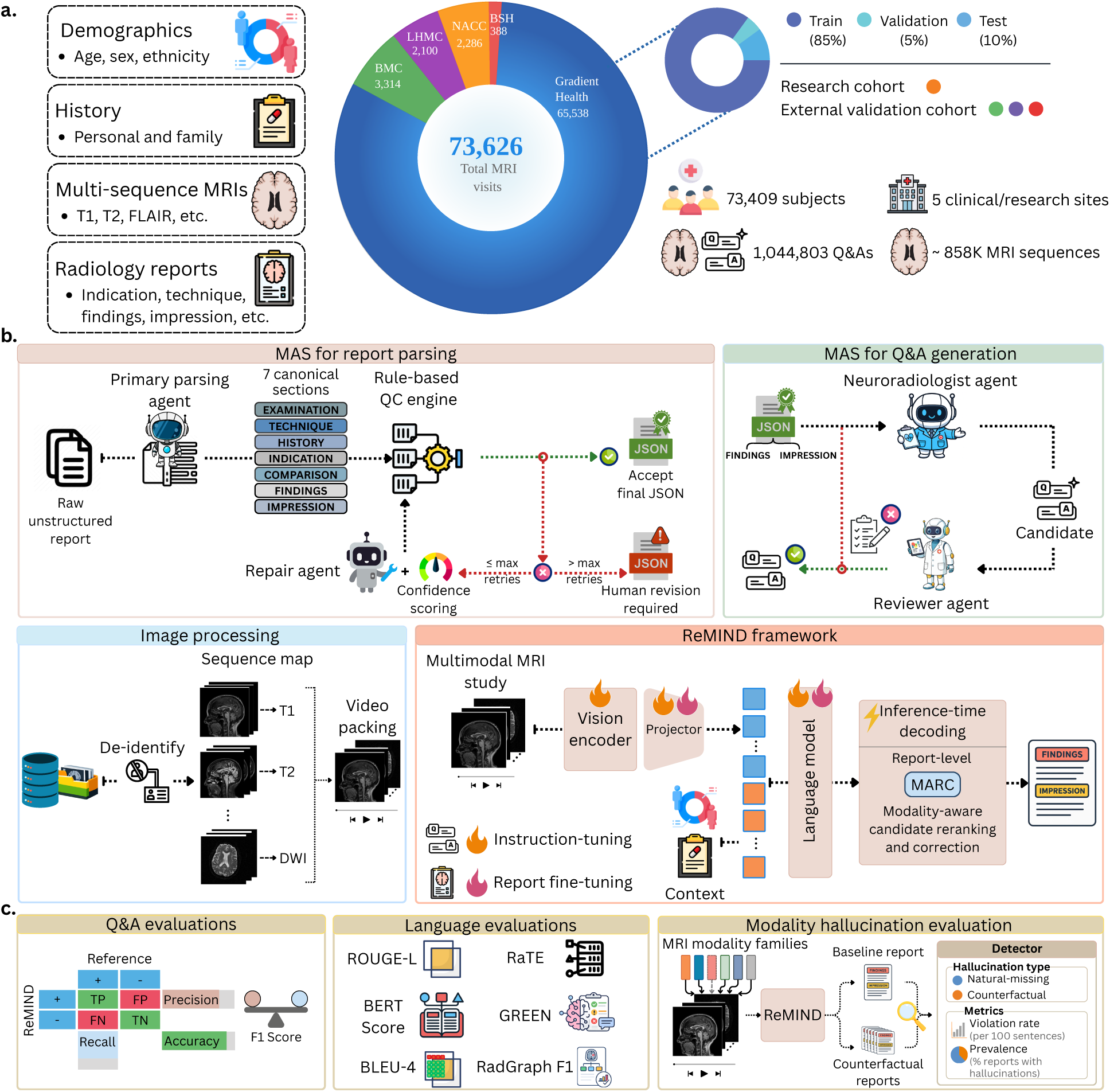
ReMIND framework. (a) Our modeling framework was developed on a large dataset comprising 73,626 multi-sequence MRI visits from 72,409 subjects across five cohorts, integrating demographics, medical history, multisequence MRI (T1, T2, FLAIR, etc.), and radiology reports. (b) Radiology reports were parsed using a multi-agent system comprising sequential large language model (LLM) agents for parsing, repair and confidence scoring, coordinated by a rule-based quality-control engine. A second multi-agent system generated question–answer pairs through interaction between neuroradiologist and reviewer agents. MRI scans were de-identified, sequence-mapped and video-packed before multimodal encoding by the ReMIND architecture, consisting of a vision encoder, multimodal projector and language model. The model was trained through instruction tuning and report fine-tuning, and performed inference using a modality-aware decoding strategy. (c) ReMIND was evaluated on visual question-answering and report generation tasks using various metrics.

**Figure 2:**
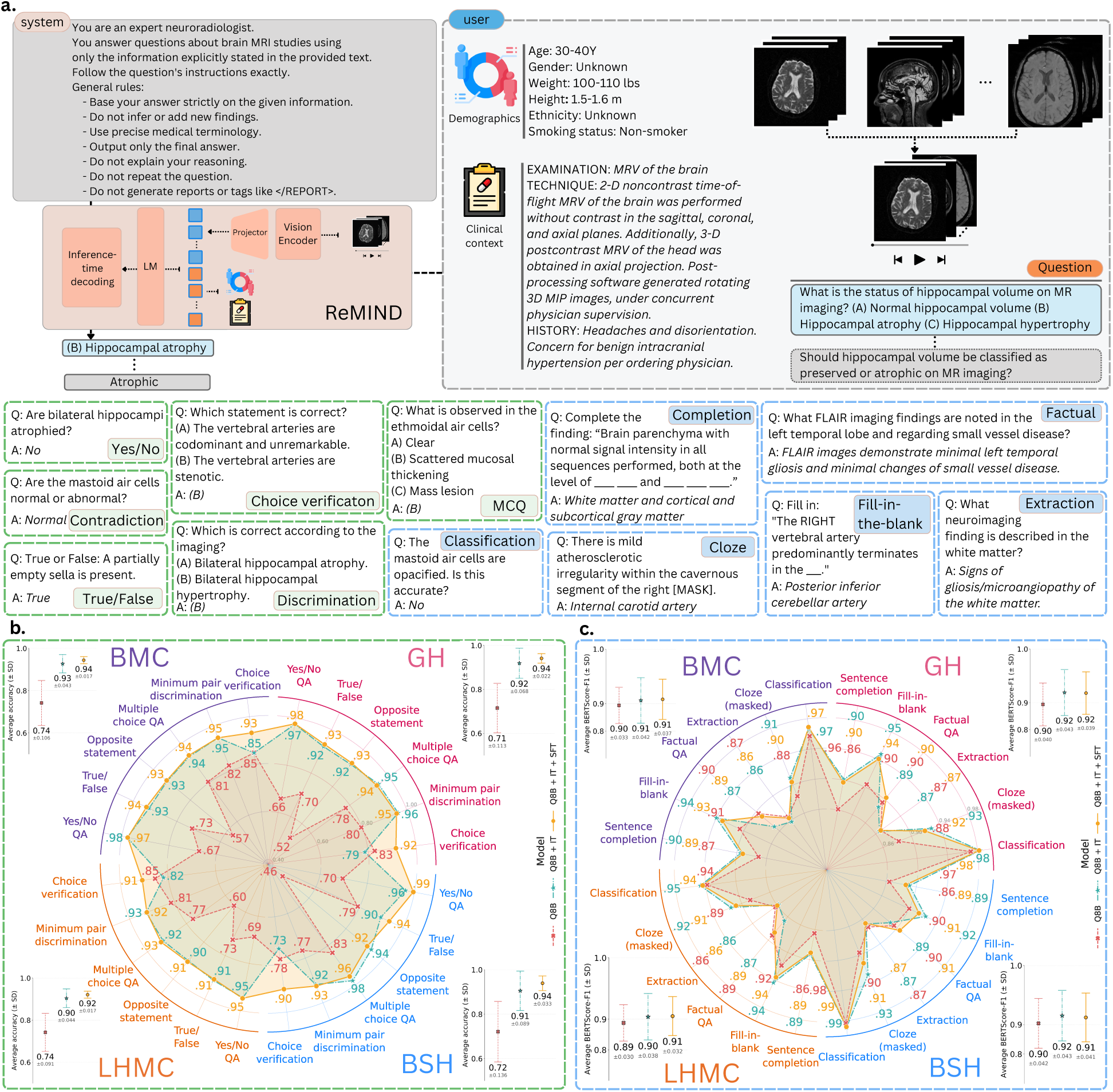
Performance of ReMIND on visual question-answering tasks. (a) Overview of the supervised fine-tuning framework employed to train ReMIND for visual question-answering tasks. Each training sample comprised a system prompt defining ReMIND’s role as a neuroradiologist, a structured clinical context, multi-sequence MRI(s), and a task-specific question-answer pair. Twelve question types were derived from radiology reports in the held-out test sets, spanning binary classification and descriptive response formats. (b) Classification accuracy across six question types requiring single-word or categorical responses. Performance is shown as a radar plot in which each quadrant corresponds to one dataset (BMC: Boston Medical Center, GH: Gradient Health, LHMC: Lahey Hospital & Medical Center, and BSH: Brain and Spine Hospital); the axes represent individual question types. Per-dataset mean accuracy with error bars is shown in the adjacent dot plot. (c) BERT-F1 score for six question types requiring sentence-level or free-text responses, displayed using the same layout as (b).

**Figure 3:**
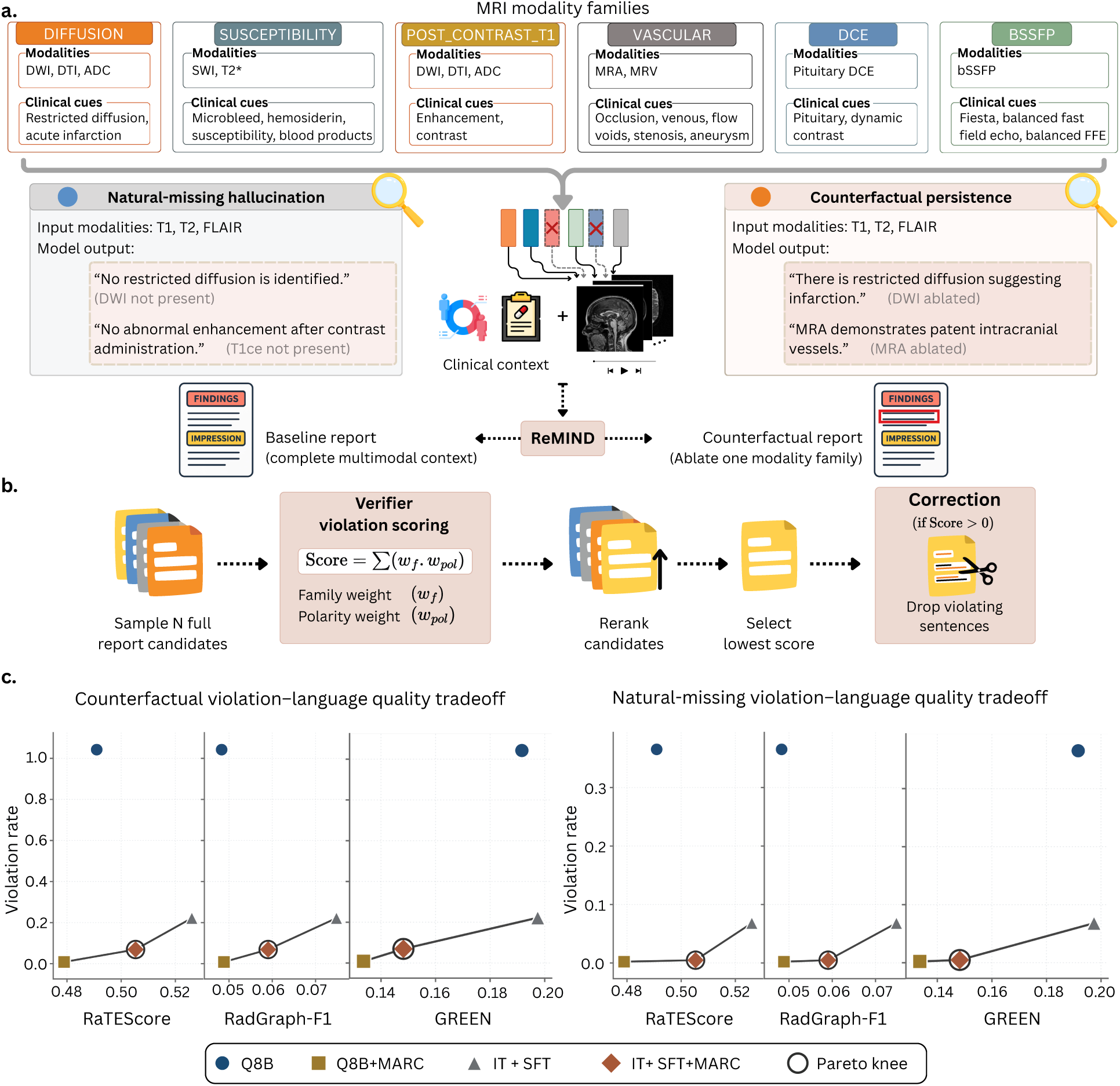
Modality hallucination detection and mitigation. (a) MRI modality families and associated clinical cue lexicon used to define modality-specific evidence constraints. Two hallucination types targeted by our framework are illustrated: natural-missing hallucination, where the model references absent modalities, and counterfactual persistence, where findings persist despite targeted ablation of modality families. (b) Overview of modality-aware candidate reranking and correction, which operates at the report level by sampling candidate reports, scoring modality violations using a verifier with family and polarity weights, reranking candidates and applying post hoc correction by dropping violating sentences. (c) trade-off between hallucination reduction (quantified as violation rate) and language quality (measured via RaTEScore, RadGraph-F1 and GREEN) under counterfactual and natural-missing settings is shown. The Pareto frontier is shown as a solid line, with points on the frontier representing non-dominated trade-offs. The circled point denotes the knee point of the Pareto frontier.

**Table 1:**
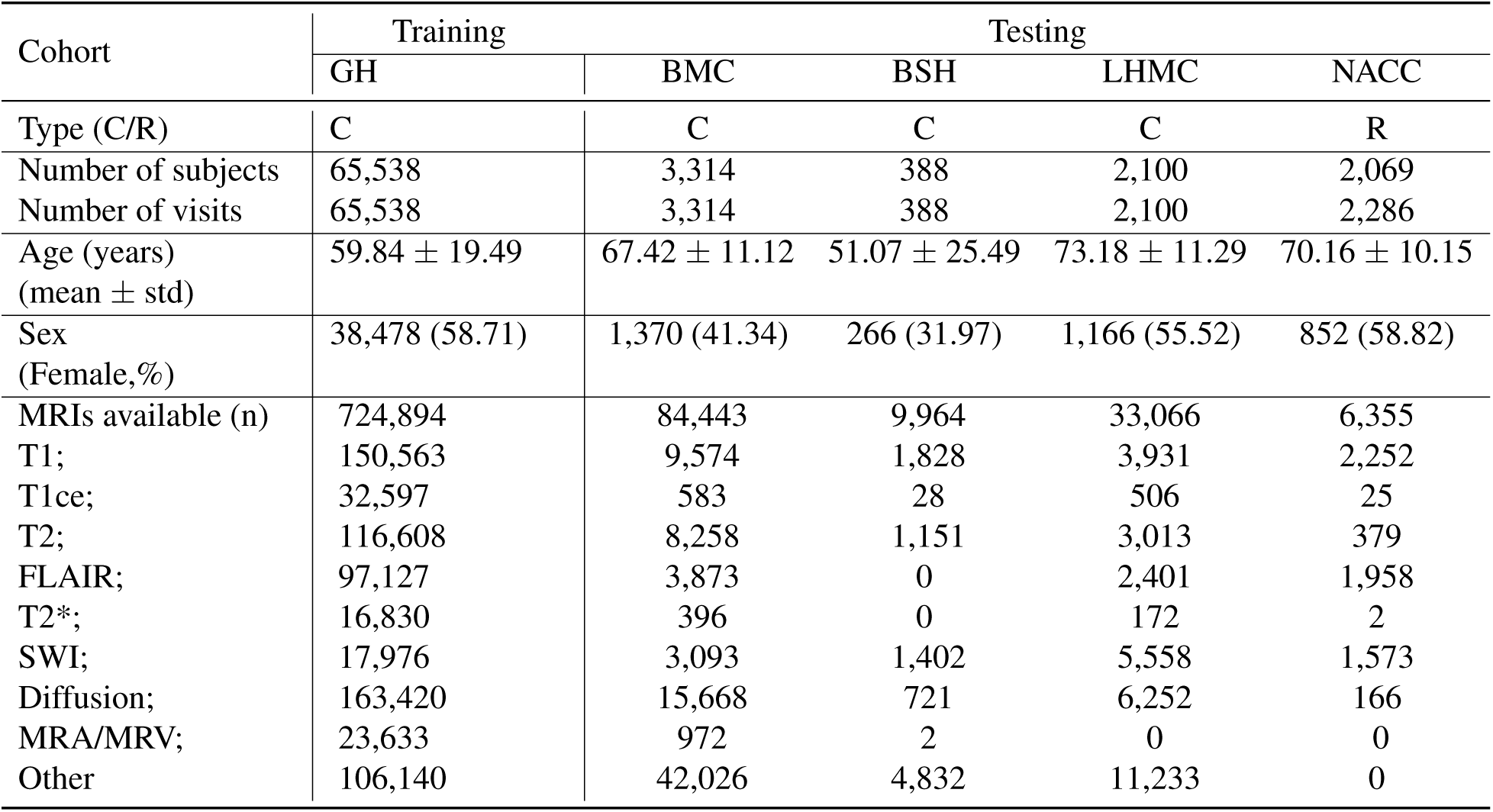
Study population. We included participants from one large training cohort (Gradient Health, GH) and four external testing cohorts (Boston Medical Center [BMC], Brain and Spine Hospital [BSH], Lahey Hospital & Medical Center [LHMC], and the National Alzheimer’s Coordinating Center [NACC]). Cohorts were either clinical (GH, BMC, BSH and LHMC) or research (NACC) in nature. Demographic summary data comprised the number of subjects and visits, mean age with standard deviation, and the percentage of female participants (with 831 patients from BMC lacking sex information). MRI sequence availability was reported as total counts by type (including T1, T1 post-contrast, T2, FLAIR, T2*/SWI, diffusionweighted, MRA/MRV, and a category for other sequences such as reformats, field maps, dynamic contrastenhanced pituitary studies, proton-density weighted sequences, balanced steady-state free precession MRIs, and unknown diagnostic scans).

**Table 2:**
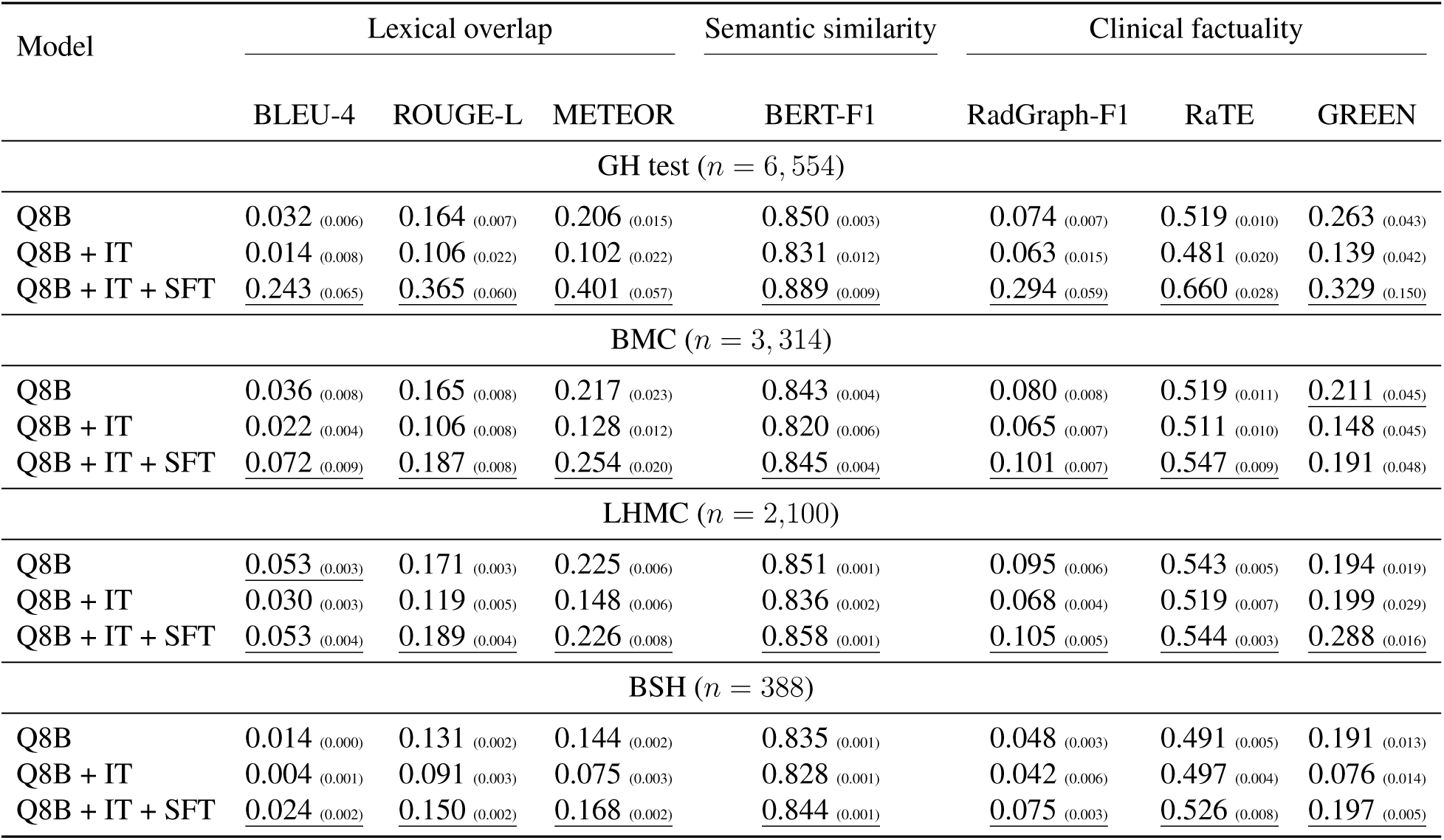
Performance on radiology report generation. Results are shown for the baseline `Q8B` model, after instruction tuning (`Q8B + IT`), and after report-specific supervised fine-tuning (`Q8B + IT + SFT`) on radiology report generation (RRG) using multiple datasets: Gradient Health test set (GH test, n = 6,554), Boston Medical Center (BMC, n = 3,314), Lahey Hospital & Medical Center (LHMC, n = 2,100), and Brain and Spine Hospital (BSH, n = 388). Metrics are grouped into three categories: (i) lexical overlap (BLEU4, ROUGE-L, METEOR), (ii) semantic similarity (BERT-F1), and (iii) clinical factuality (RadGraph-F1, RaTE, GREEN), with all values reported as higher-is-better (*↑*). Standard deviations are shown in subscript immediately following each metric value, and the best performance in each column within each cohort is underlined.

| Model | Lexical overlap |  |  | Semantic similarity | Clinical factuality |  |  |
| --- | --- | --- | --- | --- | --- | --- | --- |
|  | BLEU-4 | ROUGE-L | METEOR | BERT-F1 | RadGraph-F1 | RaTE | GREEN |
| GH test ($n = 6,554$) | | | | | | | |
| Q8B | 0.032<sub>(0.006)</sub> | 0.164<sub>(0.007)</sub> | 0.206<sub>(0.015)</sub> | 0.850<sub>(0.003)</sub> | 0.074<sub>(0.007)</sub> | 0.519<sub>(0.010)</sub> | 0.263<sub>(0.043)</sub> |
| Q8B + IT | 0.014<sub>(0.008)</sub> | 0.106<sub>(0.022)</sub> | 0.102<sub>(0.022)</sub> | 0.831<sub>(0.012)</sub> | 0.063<sub>(0.015)</sub> | 0.481<sub>(0.020)</sub> | 0.139<sub>(0.042)</sub> |
| Q8B + IT + SFT | <u>0.243</u><sub>(0.065)</sub> | <u>0.365</u><sub>(0.060)</sub> | <u>0.401</u><sub>(0.057)</sub> | <u>0.889</u><sub>(0.009)</sub> | <u>0.294</u><sub>(0.059)</sub> | <u>0.660</u><sub>(0.028)</sub> | <u>0.329</u><sub>(0.150)</sub> |
| BMC ($n = 3,314$) | | | | | | | |
| Q8B | 0.036<sub>(0.008)</sub> | 0.165<sub>(0.008)</sub> | 0.217<sub>(0.023)</sub> | 0.843<sub>(0.004)</sub> | 0.080<sub>(0.008)</sub> | 0.519<sub>(0.011)</sub> | <u>0.211</u><sub>(0.045)</sub> |
| Q8B + IT | 0.022<sub>(0.004)</sub> | 0.106<sub>(0.008)</sub> | 0.128<sub>(0.012)</sub> | 0.820<sub>(0.006)</sub> | 0.065<sub>(0.007)</sub> | 0.511<sub>(0.010)</sub> | 0.148<sub>(0.045)</sub> |
| Q8B + IT + SFT | <u>0.072</u><sub>(0.009)</sub> | <u>0.187</u><sub>(0.008)</sub> | <u>0.254</u><sub>(0.020)</sub> | <u>0.845</u><sub>(0.004)</sub> | <u>0.101</u><sub>(0.007)</sub> | <u>0.547</u><sub>(0.009)</sub> | 0.191<sub>(0.048)</sub> |
| LHMC ($n = 2,100$) | | | | | | | |
| Q8B | <u>0.053</u><sub>(0.003)</sub> | 0.171<sub>(0.003)</sub> | 0.225<sub>(0.006)</sub> | 0.851<sub>(0.001)</sub> | 0.095<sub>(0.006)</sub> | 0.543<sub>(0.005)</sub> | 0.194<sub>(0.019)</sub> |
| Q8B + IT | 0.030<sub>(0.003)</sub> | 0.119<sub>(0.005)</sub> | 0.148<sub>(0.006)</sub> | 0.836<sub>(0.002)</sub> | 0.068<sub>(0.004)</sub> | 0.519<sub>(0.007)</sub> | 0.199<sub>(0.029)</sub> |
| Q8B + IT + SFT | <u>0.053</u><sub>(0.004)</sub> | <u>0.189</u><sub>(0.004)</sub> | <u>0.226</u><sub>(0.008)</sub> | <u>0.858</u><sub>(0.001)</sub> | <u>0.105</u><sub>(0.005)</sub> | <u>0.544</u><sub>(0.003)</sub> | <u>0.288</u><sub>(0.016)</sub> |
| BSH ($n = 388$) | | | | | | | |
| Q8B | 0.014<sub>(0.000)</sub> | 0.131<sub>(0.002)</sub> | 0.144<sub>(0.002)</sub> | 0.835<sub>(0.001)</sub> | 0.048<sub>(0.003)</sub> | 0.491<sub>(0.005)</sub> | 0.191<sub>(0.013)</sub> |
| Q8B + IT | 0.004<sub>(0.001)</sub> | 0.091<sub>(0.003)</sub> | 0.075<sub>(0.003)</sub> | 0.828<sub>(0.001)</sub> | 0.042<sub>(0.006)</sub> | 0.497<sub>(0.004)</sub> | 0.076<sub>(0.014)</sub> |
| Q8B + IT + SFT | <u>0.024</u><sub>(0.002)</sub> | <u>0.150</u><sub>(0.002)</sub> | <u>0.168</u><sub>(0.002)</sub> | <u>0.844</u><sub>(0.001)</sub> | <u>0.075</u><sub>(0.003)</sub> | <u>0.526</u><sub>(0.008)</sub> | <u>0.197</u><sub>(0.005)</sub> |

### Performance on visual question-answering tasks

We first evaluated ReMIND across VQA benchmarks derived from radiological reports on the clinical datasets, spanning six discriminative classification and six open-ended free-form generative question types (Fig. 2a). For classification tasks (Fig 2b), the base model (`Q8B`) exhibited higher performance variability across all datasets and was near chance on contradiction and yes/no questions. Instruction tuning (`Q8B+IT`) reduced this variability by improving performance on the weakest question types, particularly contradiction questions (accuracy gains of 49.42 *−* 104.59%) and binary verification tasks (yes/no: 37.70 *−* 47.80%; true/false: 13.38 *−* 30.78%). Supervised report finetuning (`Q8B+IT+SFT`) further reduced cross-type variability on every dataset. Across all six classification question types, `Q8B+IT+SFT` achieved the highest average accuracy gains on every dataset (GH: 31.61%; BMC: 27.22%; LHMC: 24.12%; BSH: 30.24%) with corresponding improvements in precision and recall across question types (Table S4).

For free-form generative tasks, ReMIND’s improvements in BERTScore-F1 were more comparatively modest, as the base model already demonstrated reasonable semantic similarity to reference answers (average BERTScore-F1: 0.894 *−* 0.902 across datasets; Fig. 2c), with low cross-type variability. Nevertheless, `Q8B+IT` yielded the largest gains on constrained completion types: cloze (2.94 *−* 5.22%), fill-in-the-blank (2.72 *−* 4.87%), and sentence completion (2.76 *−* 4.65%), but degraded factual QA performance across all sites, reflecting a shift toward more verbose and paraphrased responses that diverge semantically from the precise reference answers. Supervised report fine-tuning (`Q8B+IT+SFT`) reversed this degradation, surpassing `Q8B` performance on factual QA at GH and BSH (gains: 0.89% and 1.12%), while preserving gains on structured question types. Overall, BERTScore-F1 improvements were comparable between `Q8B+IT+SFT` and `Q8B+IT` (`Q8B+IT+SFT`: 1.11–2.34%; `Q8B+IT`: 1.00–2.45%), with neither variant consistently dominant. However, `Q8B+IT+SFT`’s key advantage lies in its more balanced profile, maintaining factual precision alongside type-level consistency across generative question types. Cross-site differences in average accuracy were small and consistent (within 2–3 percentage points for equivalent models), indicating that the multi-agent pipeline generated benchmarks of comparable difficulty across datasets despite differences in report style and patient demographics; this pattern was also reflected in the per-type lexical overlap metrics across generative question types (Table S5).

To assess ReMIND’s ability to integrate complementary information across MRI sequences, we further evaluated it on a targeted multi-choice VQA benchmark constructed from NACC cases with documented imaging evidence (Table S6). The benchmark leveraged structured annotations from key NACC variables to generate questions that implicitly required cross-sequence reasoning, drawing on complementary contrasts such as FLAIR for white-matter changes, SWI for microbleeds, T1 for atrophy, and DWI/ADC for acute ischemia, without explicitly indicating relevant sequences. Instruction-tuned model (`Q8B+IT`) achieved balanced and competitive results, with notable strengths in infarcts and atrophy, where multi-sequence input appeared to reduce both missed positives and overconfident false alarms. Across all categories, the model benefited noticeably from complementary contrasts, improving detection of subtle or sequence-specific findings that single-modality approaches often overlooked. These gains in balanced precision–recall trade-offs underscore the value of domain-specific multi-contrast training, suggesting that carefully tailored models can deliver clinically meaningful performance without relying on frontier-scale resources.

### Impact of instruction tuning and fine-tuning on report generation

Instruction tuning alone (`Q8B+IT`) reduced the performance on radiology report generation (RRG) across all metric categories (Table 2). On the GH test set, this was reflected by reductions in GREEN (0.26 *→* 0.14), RaTEScore (0.52 *→* 0.48), and BERT-F1 (0.85 *→* 0.83), indicating that general-purpose instruction alignment alone may be insufficient for structured RRG. In contrast, subsequent report-specific fine-tuning, as implemented in `Q8B+IT+SFT`, led to improvements across all metrics. On GH, `Q8B+IT+SFT` increased lexical overlap (ROUGE-L: 0.16 *→* 0.37), semantic similarity (BERT-F1: 0.85 *→* 0.89), and clinical factuality (RadGraph-F1: 0.07 *→* 0.30; GREEN: 0.26 *→* 0.38), demonstrating strong gains in linguistic and clinically grounded performance. These improvements generalized across external datasets, albeit with reduced magnitude. On BMC and LHMC, `Q8B+IT+SFT` consistently improved clinical factuality metrics relative to both `Q8B` and `Q8B+IT`, with gains in RadGraph-F1 and GREEN, indicating improved structured reasoning and entity-level correctness under distribution shift. On the smaller and more challenging BSH cohort comprising brain tumor cases, `Q8B+IT+SFT` restored performance degraded by instruction tuning and achieved the best overall balance across metrics, including GREEN (0.20 vs. 0.19 for `Q8B` and 0.08 for `Q8B+IT`). Overall, these results demonstrate that while instruction tuning alone may misalign the model with the structured and domain-specific nature of radiology reporting, subsequent report-level fine-tuning, as realized in `Q8B+IT+SFT`, is important to recover and improve linguistic quality and clinical factuality, with consistent benefits across in-domain and external datasets.

### Trade-off between hallucination reduction and report quality

We examined the relationship between hallucination reduction and language quality across decoding strategies (Fig. 3a & Fig. 3b). Each point represents a model-decoding configuration evaluated under natural-missing and counterfactual settings (Fig. 3c), with report quality measured by RaTEScore,^18^ RadGraph-F1,^19^ GREEN ^20^ (x-axis) and hallucination quantified using violation rate (y-axis). Under greedy decoding, both the base model (`Q8B`) and `Q8B+IT+SFT` achieved strong language quality across all metrics, with `Q8B+IT+SFT` outperforming Q8B on all three metrics. However, both models exhibited hallucination rates, indicating frequent unsupported modality attributions despite improved fluency and alignment. Applying modality-aware candidate reranking and correction (MARC) reduced violation rates across counterfactual and natural-missing settings for both checkpoints. This reduction was accompanied by a shift toward lower language quality across the three metrics, reflecting a trade-off between hallucination suppression and report quality. The Pareto frontier exhibited a knee point, corresponding to a balanced trade-off between these objectives.

Across all evaluation metrics, the same qualitative trend was observed: MARC suppressed hallucinations regardless of the underlying model. When applied to the pretrained `Q8B` model, MARC produced a larger drop in RaTEScore, RadGraph-F1 and GREEN, corresponding to a more conservative operating regime. In contrast, when applied to `Q8B+IT+SFT`, MARC achieved comparable reductions in violation rate while preserving higher language quality. These results indicate that MARC interacts with model alignment, yielding different trade-off profiles depending on the degree of instruction tuning and domain adaptation. Improvements observed from report-specific fine-tuning (Table 2) were complementary to the gains achieved through modality-aware decoding (Fig. 3). While `Q8B+IT+SFT` improved lexical, semantic and clinically grounded agreement with reference reports, it does not explicitly enforce consistency with the available input modalities. As shown in Fig. 3, hallucination errors persisted even in `Q8B+IT+SFT` under standard decoding. In contrast, MARC operated at inference time to enforce modality consistency, reducing unsupported modality attributions without requiring additional training. Together, these results highlight two orthogonal axes of improvement: model training, which enhances report quality and clinical alignment, and decoding-time control, which enforces input-faithful generation.

## Discussion

ReMIND represents a targeted advance in multimodal AI for neuroimaging. Trained on a large-scale dataset of multi-sequence brain MRIs paired with diverse question–answer annotations and radiologist reports, the framework generates clinically relevant, structured, and reliable radiology reports. Through extensive training, ReMIND acquired comprehensive domain knowledge of neurological disorders, enabling strong performance in medical visual question answering and radiology report generation across various cohorts.

ReMIND’s performance stems from an orchestrated pipeline that balanced domain adaptation with clinical alignment. Large-scale instruction tuning on over one million grounded question–answer pairs provided the model with robust domain knowledge and multimodal understanding, enabling it to interpret complex neuroimaging patterns in a manner consistent with expert neuroradiological thinking. Subsequent report-specific supervised fine-tuning then augmented its capability towards a structured, concise and factually constrained language required for radiology reporting, yielding gains in lexical alignment, semantic coherence and entity-level correctness. The addition of modality-aware candidate reranking and correction (MARC) at inference time introduced a final layer of fidelity: by evaluating complete report candidates against a violation-weighted modality cue lexicon and selectively correcting or suppressing unsupported statements, MARC ensured that generated text remained bound to the actual input sequences. Together, these components form a complementary system that progressively refines the model’s internal representations and its outward trustworthiness, demonstrating that targeted engineering can deliver reliable, deployable performance in high-stakes medical imaging without requiring frontier-scale resources. The design of ReMIND also highlights a broader principle in medical AI development: specialization can outperform general-purpose scale in domains where factual grounding and safety are paramount. By prioritizing domain-specific tuning over massive parameter counts, the framework achieves high reliability while remaining computationally accessible. This approach not only reduces the barriers to deployment in resource-limited settings but also offers a model for building trustworthy tools in other high-stakes medical imaging tasks, where hallucinations or unsubstantiated claims carry direct consequences for patient care.

Our study has some limitations. While ReMIND was trained and validated on a large, diverse set of brain MRI studies, the dataset primarily reflects adult populations from clinical and research settings, with negligible representation of pediatric cases and certain underrepresented racial or ethnic groups. This may restrict generalizability to broader global populations or specific demographic subgroups where disease presentation or imaging protocols differ. Coverage of neurological disorders is also incomplete; although the model was exposed to a wide range of neurodegenerative, vascular, inflammatory, and neoplastic conditions, rarer entities and acute pediatric pathologies remain underrepresented. Finally, while MARC reduced modality-specific hallucinations, residual errors persist in some edge cases, and the framework has not yet been exhaustively tested across the full spectrum of scanner vendors, field strengths, acquisition protocols, or post-processing variations encountered in routine practice.

In conclusion, ReMIND demonstrates that targeted, domain-specific modeling can achieve clinically meaningful performance in multi-sequence brain MRI interpretation without relying on frontier-scale resources. By combining large-scale instruction tuning, report-specific fine-tuning, and inference-time faithfulness constraints, ReMIND produced reliable outputs across diverse cohorts. Our findings establish a practical pathway for AI to support consistent, equitable neuroimaging evaluation, directly addressing realworld diagnostic challenges in neurological care.

## Methods

The study relied on secondary analysis of de-identified electronic medical records and neuroimaging data from clinical and research cohorts sourced from various institutions. These institutions had previously obtained ethical approval for data collection and sharing, and the secondary use of these datasets for model training, fine-tuning, and evaluation was reviewed and approved by the relevant institutional review boards or ethics committees. All processing adhered to de-identification standards and applicable privacy regulations to ensure full compliance with ethical and regulatory requirements.

### Study population

Our study included five cohorts: a large clinical cohort for model training (Gradient Health [GH], *n* = 65,538 visits) and four external testing cohorts (Boston Medical Center, Boston, MA, USA [BMC], *n* = 3,314; Lahey Hospital & Medical Center, Burlington, MA, USA [LHMC], *n* = 2,100; Brain and Spine Hospital, Chennai, India [BSH], *n* = 388; and National Alzheimer’s Coordinating Center [NACC; research cohort], *n* = 2,286 visits ^21^). Prediction and evaluation were performed at the MRI visit level (an imaging study linked to its radiology report), generating 73,626 visits from 73,409 unique subjects (Table1; Fig. 1a). All clinical cohorts provided participant-level demographics and comprehensive multi-sequence brain MRI studies, each paired with a radiology report. These reports were parsed and mapped to seven canonical sections for consistency: EXAMINATION, TECHNIQUE, COMPARISON, HISTORY, INDICATION, FINDINGS and IMPRESSION. For the NACC cohort, the selection was based on the availability of structured neuroimaging evidence in the Uniform Data Set.

### Selection criterion

We included brain MRI studies (hereafter referred to as MRI), comprising conventional sequences (e.g., T1-weighted, T2-weighted, FLAIR, diffusion-weighted, etc.) and, when acquired during the same visit, angiographic sequences (MRA/MRV). Inclusion was guided by institution-specific criteria to ensure anatomical relevance, interpretive completeness and clinical utility for neurological disorders.

For the GH cohort, visits were identified through a deterministic rule-based screening pipeline applied to DICOM metadata and linked radiology report text. Examinations were first required to have the `BodyPartExamined` field consistent with head or brain imaging, with accommodation for institutionspecific variants and multi-valued entries. Reports were then screened for interpretive completeness: visits were excluded if the report indicated that interpretation would be provided separately, unless a clearly delineated IMPRESSION section was present. Anatomical relevance was further refined by screening the DICOM `StudyDescription` field against predefined inclusion and exclusion term sets (Table S1). Examinations were retained if the description included intracranial indicators (e.g., brain, head, cranial, pituitary/sellar, or angiography-related terms such as MRA/MRV) and excluded if it suggested non-brain-only protocols (e.g., spine-only or extracranial head/neck indications).

For the external clinical cohorts (BMC, LHMC and BSH), cases were collected under separate institutional review board (IRB)-approved protocols authorizing extraction of de-identified comprehensive MRI studies, patient-level demographics (i.e., age, sex, ethnicity, clinical history), and associated radiology reports. Exclusion criteria included absence of an MRI study, lack of a linked radiology report, or failure to satisfy keyword-based search criteria for neurological relevance. Where supported by the local picture archiving and communication system (PACS), additional filters restricted the dataset to MRI modality, head/brain body part, ordering services from neurology, psychiatry or memory clinic, unrestricted age, and institution-specific date ranges. Data extraction was performed via structured query language (SQL) queries on the electronic health record (EHR) system to retrieve fully de-identified datasets. All extraction activities were logged and audited to ensure traceability, regulatory compliance and prevention of unauthorized access.

Visits from the NACC cohort were selected based on the availability of structured neuroimaging evidence in the Uniform Data Set ^21^. We included cases with non-missing responses in the imaging variables CVDIMAG1 (single strategic infarct), CVDIMAG2 (multiple infarcts), HIPPATR (hippocampal atrophy), IMAGLAC (lacunar infarcts), IMAGMACH (macrohemorrhages), IMAGMICH (microhemorrhages), IMAGEWMH (extensive white-matter hyperintensity), INFNETW (cystic infarction in cognitive networks), INFWMH (combined infarction, extensive WMH, and executive impairment), STKIMAG (stroke confirmed by neuroimaging), and MRFTLD (frontal or anterior temporal atrophy suggestive of frontotemporal lobar degeneration). These fields provided reliable binary or categorical indicators of key structural abnormalities relevant to neurodegenerative and vascular pathologies.

### Image processing

Raw DICOM studies were securely transferred via encrypted channels (e.g., SFTP with end-to-end encryption) to dedicated research computing servers at Boston University, hosted within a university-managed secure environment. Access was restricted to authorized personnel through multifactor authentication and role-based controls, with versioning and regular backups implemented to safeguard data integrity and prevent loss. Subsequent processing, including de-identification, quality control, and format conversion, was performed within this secure environment. All data were processed at the visit level, originating from de-identified DICOM studies containing one or more MRI sequences (Table 1). For cohorts requiring additional de-identification (LHMC, BMC and BSH), DICOM headers were automatically scanned and redacted in accordance with HIPAA guidelines. Sensitive fields, including acquisition dates and site-specific identifiers, were removed or generalized (e.g., dates shifted to relative offsets such as “Day 0”), while technically relevant imaging parameters such as voxel spacing, slice thickness, magnetic field strength, and echo/repetition times (when present), were preserved to support downstream analysis and auditability.

DICOM series were converted to NIfTI format using an automated, study-level pipeline built around the `dcm2niix` package, with parallelized execution across cases to support large-scale processing. Individual series were first extracted from compressed archives and organized at the study and series levels prior to conversion. Conversion was performed using standardized parameters, including gzip compression and consistent filename templating to preserve acquisition metadata (e.g., protocol name, series number, and acquisition time). A fallback decompression step using the GDCM toolkit (`gdcmconv`) was applied when initial conversion failed, enabling recovery of non-standard or compressed DICOM encodings. Postconversion automated validation checks, including verification of non-empty outputs and consistency of image dimensions, confirmed successful NIfTI generation. Series that failed conversion or produced empty outputs, due to corrupted files, missing slices, or inconsistent dimensions, were automatically discarded by the conversion pipeline. Finally, a subset of *n*=100 randomly selected studies underwent manual review that validated the automated quality-control outputs, ensuring consistent, analysis-ready inputs for downstream model training and evaluation.

### Rule-based MRI sequence labeling

To standardize series across heterogeneous naming conventions used by different institutions and scanner vendors, we developed a deterministic rule-based pipeline that maps each MR series to one of the following canonical labels (Fig. 1a): T1, post-contrast T1 (T1CE), T2, FLAIR, susceptibility-weighted imaging (SWI), T2*, diffusion-derived series (DWI, ADC, DTI), and angiographic series (MRA, MRV). Labels were inferred from the series filename and any available sidecar metadata, including `SeriesDescription`, `ProtocolName`, `ScanOptions`, `ImageType`, `BidsGuess`, and `EchoTime`. The pipeline first excluded known duplicate or equalized volumes based on filename suffixes, then identified diffusion acquisitions via the presence of `.bval` and `.bvec` files, classifying them as DTI when non-trivial gradient directions were present and as DWI otherwise. Subsequently, a hierarchical pattern-matching procedure was applied to the metadata text to distinguish localizers and reformats from diagnostic sequences, while additional deterministic rules detected field maps (using vendor-specific signatures based on echo time and gradient-echo image types), pituitary dynamic contrast-enhanced studies, and MR venography (Table S2). Non-diagnostic series were excluded from model inputs: specifically, series labeled `LOCALIZER` (e.g., scout or survey acquisitions) and `OTHER` (including processed images and non-diagnostic exports) were discarded. Series labeled `REFORMAT` were retained, however, because for a subset of visits (n=53,901 from GH; n=11,196 from LHMC), diagnostic image content was available only as reconstructed or reformatted series rather than the original acquisitions.

### Volume normalization and video packing

To prepare multi-sequence MRI data for VLM input, each 3D series was converted into a fixed-length sequence of 2D frames through the video-packing pipeline (Fig. 1b). Volumes were loaded as grayscale arrays and resampled in-plane to a uniform spatial resolution of *H* = *W* = 128 pixels using linear interpolation, while preserving the original through-plane dimension. For 4D time-series acquisitions, each 3D subvolume was processed independently following the same steps. Per-volume intensity normalization was performed using min-max scaling, followed by linear mapping to an 8bit grayscale range (0-255). Volumes exhibiting a degenerate (constant) intensity range were excluded from further use. To avoid enforcing a fixed anatomical orientation while respecting acquisition geometry, the primary slice direction was inferred from the DICOM header’s voxel spacing: the axis with the largest voxel spacing (typically the through-plane direction) was identified as depth. The volume array was permuted accordingly and forced into a canonical tensor shape of [1*, D, H, W*] prior to frame extraction. Frames were then sampled uniformly along this depth dimension, selecting up to *K* = 256 evenly spaced slices across the full extent of the volume. To exclude background-only content, all-zero slices were removed before sampling; any volume that yielded no remaining non-zero slices was discarded. Each retained slice was converted to a single-channel image. For VLM input, all sampled frames from a given visit were concatenated into a single contiguous video-style sequence and embedded as a unified visual block within the user message. When multiple volumes existed for the same modality within a visit, each was normalized and sampled independently, with their frames appended sequentially to form the final video input.

### Report parsing

Radiology reports were processed at the visit level and mapped to seven canonical sections (EXAMINATION, TECHNIQUE, COMPARISON, HISTORY, INDICATION, FINDINGS and IMPRESSION) to enable supervised fine-tuning and model evaluation across clinical documentation formats (Fig. 1b). All report processing was performed on de-identified text. We employed an LLM-based multiagent parsing pipeline using `Qwen3-Next-80B-A3B-Instruct` to segment each GH report into the seven target sections noted above (Fig. 1b). The model was instructed to perform faithful extraction (not rewriting), preserve all clinical meaning and dates, and apply only minimal formatting cleanup (e.g., removal of section headers, administrative workflow statements, duplicated text and de-identification artifacts). Parsing was executed in asynchronous batches to maximize throughput, with a deterministic first-pass decoding configuration (temperature=0, top*−p*=1) to promote stable outputs at scale. Each parsed report was then post-processed with deterministic text normalization (e.g., removing leaked headers within section text and mapping explicit “no prior imaging/no comparison” statements to an empty COMPARISON field) and evaluated by an automated quality check (QC) engine. QC produced a binary PASS/FAIL label and a set of interpretable flags reflecting common error modes (Fig. S1). These checks included: (i) structural constraints (all seven sections present as strings; no residual header tokens within section bodies), (ii) section presence consistency (whether a section should exist based on header aliases and content cues in the raw report, with explicit exceptions when the report states “None/NA”), and (iii) grounding checks that penalize missing content and unsupported additions. Grounding was assessed using n-gram overlap and token-recall measures between the extracted section text and the raw report, together with targeted clinical “unsafe addition” checks that flag new anatomy/pathology/severity terms not supported by the source. Reports without QC flags were labeled PASS; otherwise they were labeled FAIL. For reports that failed QC, we performed iterative repair passes up to a fixed maximum number of retries. In each retry, the model was given the original report, the current parsed output, and the QC flags as guidance and instructed to correct only the problematic parts while preserving all supported content. To reduce repeated failure patterns, retry decoding used a scheduled sampling policy in which sampling became gradually less conservative across successive retries (temperature increased and top-p decreased in predefined steps). After each retry pass, we re-ran the same QC engine; the retry output was retained if it either achieved QC PASS or reduced the number of QC flags relative to the previous version. We additionally recorded per-report logs (section-level before/after changes, QC flags, and post-retry status) for auditability.

Reports from the external clinical cohorts (BMC, BSH and LHMC), were parsed into the same canonical sections using a rule-based header and boundary detection pipeline, based on curated header aliases (e.g., TECHNIQUE/PROTOCOL, HISTORY/CLINICAL HISTORY, INDICATION/REASON FOR EXAM, FINDINGS and IMPRESSION/CONCLUSION) and deterministic boundary rules, followed by minimal cleanup (removal of administrative lines and bracketed de-identification artifacts). We applied a uniform de-identification screen prior to downstream modeling to ensure that all text used in this study contained no direct identifiers. For BSH, reports received as PDFs containing the radiology report text and corresponding clinical context were first processed through an OCR python package (pytesseract) to extract the raw text. Regular expressions were then applied in sequence to identify and isolate report sections, clean OCR-introduced noise, and remove PHI. For BMC and LHMC, the transferred report text and associated fields were largely anonymized prior to transfer; we verified de-identification and removed any residual identifiers before use. In all external cohorts, this verification step targeted direct identifiers (e.g., names MRNs, addresses and contact information) and contextual identifiers (e.g., clinician names of facility references). A random subset of *n*=100 reports was manually reviewed, consistent with the audit protocol applied to imaging data, to confirm the absence of residual PHI while preserving clinically relevant content.

### Instruction tuning dataset

To provide rich and diverse supervision for instruction tuning and alignment, we generated a large-scale dataset of question–answer (QA) pairs directly from the FINDINGS and IMPRESSION sections of radiology reports in the GH cohort. This was accomplished using a multi-agent system (MAS) built on the state-of-the-art open-source language model `Qwen3-Next-80BInstruct` (Fig.1b and Fig.S2). The system consisted of a neuroradiologist agent that generated QA pairs and a neuroradiologist reviewer agent that enforced rigorous quality control. Generation followed a structured prompting framework designed to emulate expert neuroradiological reasoning (Section S2). The system prompt directed the generator to identify and characterize neurological abnormalities using specified MRI sequences while producing clinically accurate, evidence-based outputs strictly grounded in the provided report sentence, without external knowledge or speculation. For each sentence in a report, the generator was prompted to produce exactly one QA pair. The question was required to be clinically meaningful, explicitly linked to imaging findings (often referencing specific sequences), and conform to one of several predefined types: factual, yes/no, multiple-choice, classification, true/false, extraction, contradiction (opposite statement), fillin-the-blank, cloze (masked word), sentence completion, minimal pair discrimination, or choice verification (Fig. 2a). The answer was required to be concise, accurate and directly derivable from the source sentence. Three few-shot examples ensured consistent formatting and style. To maintain clinical fidelity and factual correctness, every generated QA pair was evaluated by the reviewer agent. Operating under a strict system prompt that forbade introducing new clinical facts, generating additional pairs, or inferring beyond the provided sentence, the reviewer assessed whether the pair was fully supported, directly answerable and faithful to the source text. Approved pairs were retained; otherwise, structured feedback was returned (identifying issues such as factual inconsistency, unsupported speculation, or type violation), triggering iterative regeneration with the same few-shot strategy. The revised prompt incorporated the original sentence, prior QA pair, and reviewer feedback, requiring a corrected version that remained clinically grounded, type-compliant and free of unsupported content. This cycle continued until approval or a predefined maximum number of attempts. The resulting instruction-tuning dataset comprised 1,044,803 high-quality QA pairs.

### Multi-sequence multi-choice visual question-answering benchmark

To test ReMIND’s performance on non-clinical cases, we constructed a visual question-answering (VQA) benchmark from cases in the NACC cohort with documented imaging evidence (*n* = 2, 286 visits). Each case included multiple MRI sequences paired with categorical QA annotations derived from structured NACC imaging variables (e.g., presence/absence of single strategic infarct, hippocampal atrophy, lacunar infarcts, microhemorrhages, extensive white-matter hyperintensity, macrohemorrhages, cystic infarction in cognitive networks, combined infarction with WMH and executive impairment, confirmed stroke, and frontal/anterior temporal atrophy for FTLD). For each imaging variable, five semantically equivalent question templates were defined to improve linguistic robustness (Section S3), yielding 21,975 QA instances per template. All questions were multiple-choice with the following options: Yes/No/Unknown, no relevant imaging data available. They were formulated to implicitly require integration of complementary contrasts (e.g., FLAIR for white-matter changes, SWI for microbleeds, T1 for atrophy, DWI/ADC for acute ischemia) without explicitly cueing which sequence(s) were relevant. This design mirrored real-world neuroradiological workflows, where clinicians must autonomously synthesize information across sequences to arrive at accurate interpretations.

### ReMIND modeling framework

We developed ReMIND starting from the open-weight `Qwen3-VL-8BInstruct` (or `Q8B`) vision-language model ^22^ and adapted it for multimodal brain MRI interpretation through a dual-stage tuning pipeline (Fig. 1b). We selected this 8-billion-parameter checkpoint because its unified multimodal transformer architecture natively supports autoregressive generation conditioned on interleaved text and long video sequences, which aligns with the requirements of multi-sequence 3D brain MRI interpretation. The model comprises three core components that we leveraged and adapted: (i) a vision encoder that extracts patch-level features from individual frames or video sequences, (ii) a learned projection layer that aligns visual features into the language model’s token embedding space, and (iii) a large autoregressive transformer decoder that jointly attends to textual and projected visual tokens across self-attention layers. This design enables the model to perform cross-modal reasoning, which is essential for synthesizing complementary MRI contrasts into coherent radiology-style reports or accurate answers to clinical VQAs. To handle the volumetric and multi-contrast nature of brain MRI, we implemented a video-style input representation tailored to this domain. Specifically, each 3D series was resampled and normalized, then converted into an ordered sequence of up to 256 uniformly spaced 2D slices. Frames from all available sequences within a single MRI visit were concatenated into one contiguous visual block. This strategy took advantage of the model’s built-in video understanding capability, avoiding the need for custom 3D convolutions or per-slice processing, and enforced a predictable maximum visual context length even across studies with highly variable protocol coverage.

We initialized the model using the publicly released instruction-tuned checkpoint of `Q8B`. We then conducted large-scale instruction tuning on our curated question-answer (QA) dataset, followed by supervised fine-tuning on aligned multi-sequence MRI–report pairs from the GH training cohort. After training, we evaluated the resulting model in zero-shot and fine-tuned regimes across internal and external cohorts for medical visual question-answering and automated radiology report generation.

### Modality hallucination detection and mitigation

During preliminary experiments with the baseline `Q8B` model on clinical brain MRI datasets, we observed some modality hallucinations, i.e., generated report sentences that attributed findings to specific sequences (e.g., “restricted diffusion on DWI” or “blooming susceptibility artifact on SWI”) even when those sequences were absent from the input. This issue was especially prominent in studies with incomplete or heterogeneous protocol coverage, which is a common scenario in clinical practice, and carried a risk of producing misleading or factually incorrect reports. These observations motivated the development of a systematic detection framework and practical inference-time mitigation strategies for ReMIND (Fig. 3).

We defined modality hallucination as any generated statement invoking a specific MRI sequence or modality-specific cue without corresponding support in the provided input. MRI series were grouped into six clinically meaningful modality families: DIFFUSION (DWI, ADC, DTI), SUSCEPTIBILITY (SWI, T2*), POST CONTRAST T1 (T1CE), VASCULAR (MRA, MRV), DCE (dynamic contrast-enhanced pituitary), and BSSFP (balanced steady-state free precession) (Fig. 3a). For every test case, we generated a baseline report using all available sequences, followed by counterfactual reports in which one modality family was ablated at a time. All generations used identical model checkpoints and fixed decoding parameters within each decoding configuration (e.g., greedy decoding or top-*p* sampling), ensuring that baseline and counterfactual comparisons were not confounded by changes in decoding strategy. Detection was performed at the sentence level. Reports were normalized and segmented into sentences, with coarse FINDINGS/IMPRESSION section identification using lightweight header detection. We then applied a curated, high-precision modality cue lexicon consisting of explicit sequence identifiers (“DWI”, “ADC”, “SWI”, “MRA”, etc.) and strongly associated radiological descriptors (“restricted diffusion”, “blooming susceptibility”, “post-contrast enhancement”, etc.). Family-specific constraints were enforced to limit false positives: vascular findings required explicit angiographic anchors, DCE and BSSFP required direct terminology, and post-contrast descriptions were distinguished from routine negated boilerplate. Flagged sentences received polarity labels (positive, negative, uncertain) based on negation and uncertainty cues, and were template-normalized (abstracting laterality, numbers, dates) to enable exact matching across baseline and ablated reports.

This framework enabled quantification of key hallucination patterns (Fig. 3a), including natural-missing hallucinations, where unsupported modality cues appear despite absence of the modality in the original input), and counterfactual persistence, where modality-specific cues persist after ablation of the corresponding modality family. In addition, we tracked polarity flips as a complementary consistency signal, defined as template-matched sentences that change polarity after ablation (e.g., positive *→* negative). We also flagged implausible modality attributions (e.g., hemorrhage explicitly linked to MRA) using targeted rules. The framework outputs sentence-level hallucination events, per-report counts, normalized violation rates (events per 100 baseline sentences), prevalence (proportion of reports containing at least one violation), and summaries of frequently observed hallucination templates (Table S3).

Building on this detection framework, we developed modality-aware candidate reranking and correction (MARC), which operates at the report level (Fig. 3b). While token-level logit manipulation approaches for hallucination mitigation in both general VLMs ^23,24^ and, more recently, in radiology report generation ^25^ operate at the next-token level and are therefore insensitive to sentential context and polarity, MARC evaluates complete, fluent report candidates. For each test case, we generated *N* = 4 candidate reports using top-*p* sampling (top-*p* = 0.95, temperature= 0.8), enabling controlled diversity in candidate outputs. Each candidate was evaluated using verifier violation scoring derived from the same modality family cue lexicon used in detection. Specifically, for each sentence, *s*, invoking a cue belonging to a missing modality family, *f*, a weighted violation score was accumulated as 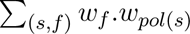.

Family weights were defined based on clinical risk and empirically observed hallucination frequency. POST CONTRAST T1 received the highest weight (*w_f_* = 2.0), reflecting the clinical implications of falsely asserting contrast-enhanced findings, followed by DIFFUSION (1.5), SUSCEPTIBILITY and VASCULAR (1.2), and DCE and BSSFP (1.0). Polarity weights reflected the asymmetric risk of different statement types, with positive assertions receiving the highest weight (*w_pol_* = 1.5), and negation and uncertainty weighted lower (1.0). Candidates were reranked according to their total violation score, and the lowest-scoring candidate was selected as the final output. When the selected candidate still carried a nonzero violation score, a sentence-level correction step was applied: violating sentences were identified and removed (up to a configurable maximum), and the remaining sentences were rejoined. If removal eliminated all content, a safe neutral fallback phrase was substituted. This correction stage was accepted only when it did not increase the violation score, preserving output quality when removal would have been counterproductive. All other generation parameters remained unchanged relative to baseline decoding.

We evaluated MARC under natural-missing and counterfactual settings, measuring hallucination reduction using violation rate and prevalence metrics, and tracking polarity changes using the detection framework. In parallel, we assessed standard radiology report generation metrics, including ROUGE ^26,27^, BERTScore ^28^, RaTEScore ^18^, RadGraph-F1 ^19^, and GREEN ^20^, to confirm that constraint gains did not compromise overall language quality. trade-off profiles were compared across all the models under MARC.

### Experimental settings

The GH cohort was reserved for model development and split into training (85%), validation (5%) and internal test (10%) sets. The training set supported large-scale instruction tuning and supervised fine-tuning; the validation set was used for hyperparameter tuning, early stopping, and checkpoint selection; and the internal GH test set served as the primary held-out evaluation for visual question answering (VQA) and radiology report generation (RRG) within the training domain. All external cohorts (BMC, BSH, LHMC and NACC) remained completely unseen during model development and were used solely for external validation of generalizability. Training was conducted for 60, 000 iterations for instruction tuning and 20, 000 iterations for report fine-tuning, with checkpoint selection based on validation performance. In both stages, the effective batch size was set to 16. In modality hallucination analyses, all generations were produced with fixed decoding parameters to prevent confounding of baseline and counterfactual comparisons by changes in decoding strategy.

### Performance metrics

We evaluated the quality of the reports using a combination of lexical overlap, semantic similarity, and clinically grounded evaluation metrics (Fig. 1c). The same lexical overlap and semantic similarity metrics were additionally applied to the open-ended generative VQA benchmarks, where model-predicted answers were compared against reference answers derived from the radiology reports. For the classification VQA benchmarks, model performance across the six discriminative question formats was assessed using accuracy, precision, and recall, capturing both overall correctness and the model’s ability to avoid false positives and false negatives across each question type. Lexical similarity between generated and reference text was measured using BLEU-4, which evaluates 4-gram precision with a brevity penalty ^29^, ROUGE-L, which measures longest common subsequence overlap between texts ^26,27^, and METEOR, which incorporates stemming and synonym matching to improve unigram alignment ^30^. To capture semantic similarity beyond surface-level token overlap, we computed BERTScore-F1, which compares contextual embeddings from pretrained Bidirectional Encoder Representations from Transformers (BERT) models to assess alignment between generated and reference texts ^28^.

Because lexical and embedding-based metrics may not fully capture the clinical correctness of radiology reports, we also employed domain-specific evaluation metrics. RaTEScore evaluates response accuracy and thematic alignment between generated and reference radiology reports ^18^. RadGraph-F1 measures factual consistency by comparing structured entity and relation graphs extracted from generated and reference reports ^19^. Finally, we report the GREEN score, a large language model–based metric designed specifically for radiology report evaluation that assesses factual correctness, completeness, and clinical relevance of generated findings ^20^. In addition to standard report quality metrics, we evaluated modality hallucination using task-specific metrics derived from our detection framework (Fig. 1c). These include the normalized violation rate (events per 100 baseline sentences) and prevalence (proportion of reports with at least one violation), enabling systematic quantification of modality-specific hallucinations under both natural-missing and counterfactual settings.

### Statistical analysis

Quantitative data were analyzed using descriptive statistics (means and standard deviations) where applicable. For VQA evaluation, standard deviations were computed at two levels: within-format, across individual samples for each question type per dataset, to assess performance consistency within categories for both classification and generative tasks; and cross-format, across the twelve question types within each dataset, to quantify homogeneity or variability across formats. These measures were calculated independently for each dataset and model variant, enabling systematic comparison of how instruction tuning and supervised fine-tuning influenced within-cohort consistency and format-level robustness. For RRG, statistics were computed across all samples within each dataset for the respective language-level metrics, providing per-cohort summaries of generation quality.

### Computational hardware and software

DICOM to NifTI conversions were performed using dcm2niix (version 1.0.20240202). Our model development utilized Python (version 3.11.12) and specifically PyTorch (version 2.8.0). We used several other Python libraries to support data analysis, including pandas (version 2.3.3), numpy (version 1.26.4), matplotlib (version 3.10.8), monai (version 1.5.1), scipy (version 1.11.4), and scikit-learn (version 1.3.2). To ensure reproducibility and avoid package conflicts across heterogeneous computing environments, all experiments were conducted within a containerized runtime using a Docker image (accessible via Singularity) based on a publicly available vision-language model environment. This setup enabled consistent dependency management and reproducible execution across the shared computing cluster. Training the model on a single Tesla H200 GPU with a shared high-performance computing cluster. The average runtime per iteration was approximately 5 seconds for instruction tuning and 12 seconds for report fine-tuning. Inference required less than one minute per MRI study. All figures were prepared using Canva and Adobe Illustrator.

## Data availability

The multi-sequence multi-choice visual question-answering (VQA) benchmark derived from the National Alzheimer’s Coordinating Center (NACC) cohort, including question-answer pairs and associated annotations, will be made publicly available on Hugging Face upon acceptance of this manuscript. Researchers interested in accessing the underlying NACC data for related studies can do so through the standard NACC data request process, which involves submitting a research proposal and agreeing to the NACC Data Use Agreement (available at https://naccdata.org/). Access to clinical cohort data can be requested from the respective institutions, subject to their IRB-approved protocols and data-sharing agreements.

## Acknowledgements

This project was supported by grants from the National Institute on Aging’s Artificial Intelligence and Technology Collaboratories (P30-AG073104 & P30-AG073105), and the National Institutes of Health (R01HL159620, R01-AG062109, R01-NS142076, R01-AG083735 and R03-DC023134).

## References

1. Scheltens, P., Fox, N., Barkhof, F. & DeCarli, C. Structural magnetic resonance imaging in the practical assessment of dementia: beyond exclusion. The Lancet Neurology 1, 13–21 (2002). URL 10.1016/S1474-4422(02)00002-9. © 2002 Elsevier Ltd. Published by Elsevier Ltd.

2. O’Brien, J. T. Role of imaging techniques in the diagnosis of dementia. British Journal of Radiology 80, S71–S77 (2007). URL 10.1259/bjr/33117326. Published: 05 March 2014.

3. Vemuri, P. & Jack Jr, C. R. Role of structural MRI in Alzheimer’s disease. Alzheimer’s Research & Therapy 2, 23 (2010). URL 10.1186/alzrt47.

4. Wardlaw, J. M. et al. Neuroimaging standards for research into small vessel disease and its contribution to ageing and neurodegeneration. Lancet Neurol 12, 822–838 (2013). STandards for ReportIng Vascular changes on nEuroimaging (STRIVE v1).

5. Filippi, M. et al. Multiple sclerosis. Nature Reviews Disease Primers 4, 43 (2018).

6. Afshari Mirak, S., Tirumani, S. H., Ramaiya, N. & Mohamed, I. The growing nationwide radiologist shortage: Current opportunities and ongoing challenges for international medical graduate radiologists. Radiology 314, e232625 (2025). URL 10.1148/radiol.232625. Published online March 4, 2025.

7. Christensen, E. W., Parikh, J. R., Drake, A. R., Rubin, E. M. & Rula, E. Y. Projected US radiologist supply, 2025 to 2055. Journal of the American College of Radiology 22, 161–169 (2025). URL 10.1016/j.jacr.2024.10.004. Accessed via Boston University.

8. Tanno, R. et al. Collaboration between clinicians and vision–language models in radiology report generation. Nature Medicine 31, 599–608 (2025). Open access. Published: 07 November 2024.

9. Bluethgen, C. et al. A vision-language foundation model for the generation of realistic chest x-ray images. Nature Biomedical Engineering 9, 494–506 (2025). Epub 2024 Aug 26.

10. Sun, Y., Wang, L., Li, G., Lin, W. & Wang, L. A foundation model for enhancing magnetic resonance images and downstream segmentation, registration and diagnostic tasks. Nature Biomedical Engineering 9, 521–538 (2025).

11. Lu, S. et al. General lightweight framework for vision foundation model supporting multi-task and multi-center medical image analysis. Nature Communications 16, 2097 (2025).

12. Chen, Q. & Hong, Y. MedBLIP: Bootstrapping language-image pretraining from 3d medical images and texts. In Cho, M., Laptev, I., Tran, D., Yao, A. & Zha, H. (eds.) Computer Vision ACCV 2024 17th Asian Conference on Computer Vision, Hanoi, Vietnam, December 8-12, 2024, Proceedings, Part III, vol. 15474 of Lecture Notes in Computer Science, 98–113 (Springer, 2024). URL 10.1007/978-981-96-0908-6_6.

13. Nath, V. et al. VILA-M3: Enhancing vision-language models with medical expert knowledge. In IEEE/CVF Conference on Computer Vision and Pattern Recognition, CVPR 2025, Nashville, TN, USA, June 11-15, 2025, 14788–14798 (Computer Vision Foundation / IEEE, 2025). URL https://openaccess.thecvf.com/content/CVPR2025/html/Nath_VILA-M3_Enhancing_Vision-Language_Models_with_Medical_Expert_Knowledge_CVPR_2025_paper.html.

14. Pan, J. et al. MedVLM-R1: Incentivizing medical reasoning capability of vision-language models (VLMs) via reinforcement learning. In International Conference on Medical Image Computing and Computer-Assisted Intervention, 337–347 (Springer, 2025).

15. Zhao, Y. et al. Med-VLM: Enhancing medical image segmentation accuracy through vision-language model. In Proceedings of the IEEE/CVF International Conference on Computer Vision (ICCV) Workshops, 7283–7293 (2025).

16. Wu, C. et al. Towards generalist foundation model for radiology by leveraging web-scale 2d&3d medical data. Nature Communications 16, 7866 (2025).

17. Zheng, S. et al. Comparison of a specialized large language model with GPT-4o for CT and MRI radiology report summarization. Radiology 316, e243774 (2025).

18. Zhao, W. et al. Ratescore: A metric for radiology report generation. In Proceedings of the 2024 Conference on Empirical Methods in Natural Language Processing, 15004–15019 (Association for Computational Linguistics, 2024). URL https://aclanthology.org/2024.emnlp-main.836/.

19. Jain, S., et al. Radgraph: Extracting clinical entities and relations from radiology reports. arXiv preprint arXiv:2106.14463 (2021).

20. Ostmeier, S. et al. Green: Generative radiology report evaluation and error notation. In Findings of the association for computational linguistics: EMNLP 2024, 374–390 (2024).

21. Beekly, D. L. et al. The national Alzheimer’s coordinating center (nacc) database: an Alzheimer disease database. Alzheimer Disease & Associated Disorders 18, 270–277 (2004).

22. Yang, A. et al. Qwen3 technical report. arXiv preprint arXiv:2505.09388 (2025). URL https://arxiv.org/abs/2505.09388.

23. Leng, S. et al. Mitigating object hallucinations in large vision-language models through visual contrastive decoding. In Proceedings of the IEEE/CVF Conference on Computer Vision and Pattern Recognition, 13872–13882 (2024).

24. Huang, Q. et al. Opera: Alleviating hallucination in multi-modal large language models via over-trust penalty and retrospection-allocation. In Proceedings of the IEEE/CVF Conference on Computer Vision and Pattern Recognition, 13418–13427 (2024).

25. Mahdavi, Z. et al. Med-vcd: Mitigating hallucination for medical large vision language models through visual contrastive decoding. Computers in Biology and Medicine 200, 111347 (2026).

26. Lin, C.-Y. & Hovy, E. Automatic evaluation of summaries using n-gram co-occurrence statistics. In Proceedings of the 2003 Human Language Technology Conference of the North American Chapter of the Association for Computational Linguistics, 150–157 (Association for Computational Linguistics, Edmonton, Canada, 2003). URL https://aclanthology.org/N03-1020.

27. Lin, C.-Y. & Och, F. J. Automatic evaluation of machine translation quality using longest common subsequence and skip-bigram statistics. In Proceedings of the 42nd Annual Meeting of the Association for Computational Linguistics (ACL-04), 605–612 (Association for Computational Linguistics, Barcelona, Spain, 2004). URL https://aclanthology.org/P04-1077.

28. Zhang, T., Kishore, V., Wu, F., Weinberger, K. Q. & Artzi, Y. Bertscore: Evaluating text generation with bert. In International Conference on Learning Representations (2020). URL https://openreview.net/forum?id=SkeHuCVFDr.

29. Papineni, K., Roukos, S., Ward, T. & Zhu, W.-J. BLEU: a method for automatic evaluation of machine translation. In Proceedings of the 40th Annual Meeting of the Association for Computational Linguistics, 311–318 (Association for Computational Linguistics, Philadelphia, Pennsylvania, USA, 2002). URL https://aclanthology.org/P02-1040.

30. Banerjee, S. & Lavie, A. METEOR: An automatic metric for MT evaluation with improved correlation with human judgments. In Proceedings of the ACL Workshop on Intrinsic and Extrinsic Evaluation Measures for Machine Translation and/or Summarization, 65–72 (Association for Computational Linguistics, Ann Arbor, Michigan, 2005). URL https://aclanthology.org/W05-0909.

