## Supplement for "Vision-language framework for multi-sequence brain magnetic resonance imaging"

### Supplementary material

#### S1 Prompt design for report parsing and repair

This section includes the exact system and user prompts for (i) primary parsing of Gradient Health (GH) radiology reports into seven canonical sections, (ii) quality-control-guided repair of failed parses, and (iii) post-repair confidence scoring. Prompts are shown verbatim with placeholders (e.g., `{report_text}`, `{parsed_json}`, `{qc_flags}`) indicating fields populated at runtime. Related workflow details are shown in Fig. S1.

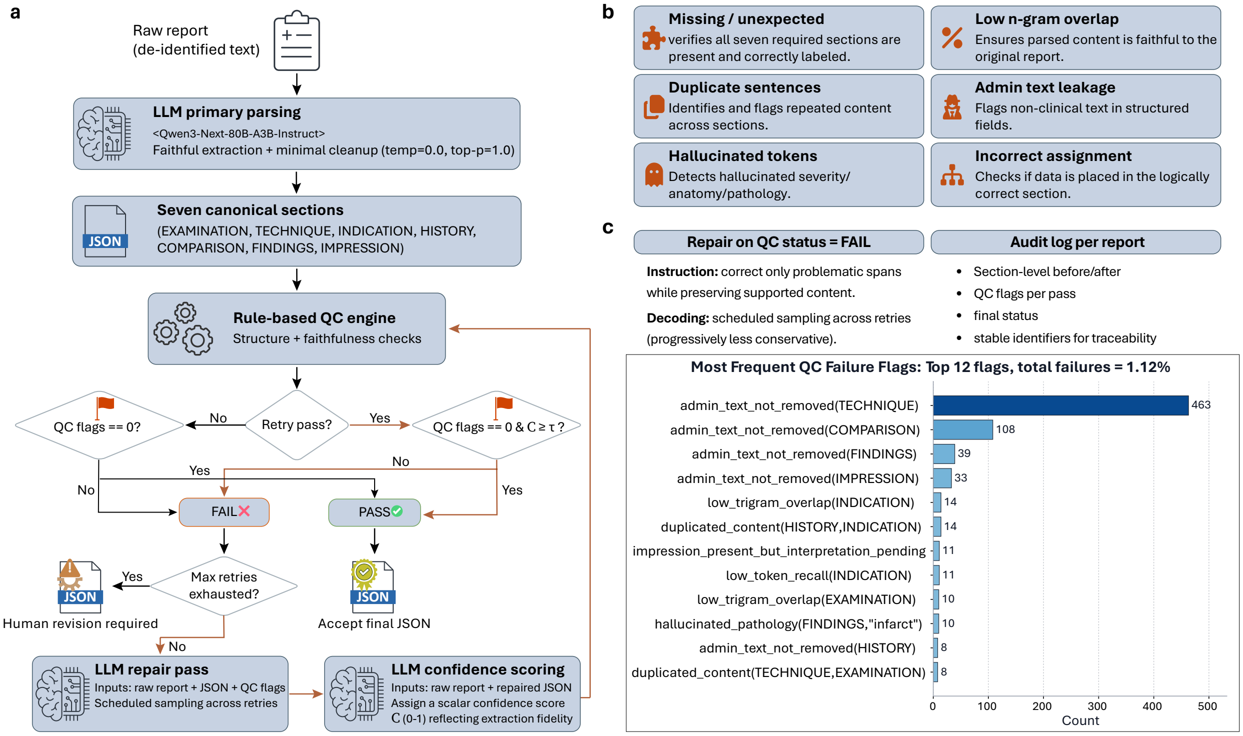


**Figure S1: Automated report parsing and quality control for Gradient Health radiology reports.** a, Overview of the LLM-based pipeline used to segment raw reports into seven canonical sections (EXAMINATION, TECHNIQUE, COMPARISON, HISTORY, INDICATION, FINDINGS, and IMPRESSION), followed by rule-based quality control (QC) and LLM retry logic, depicted by orange arrows, with a maximum number of retries ($n$=10). b, QC error modes and corresponding rule-based flags used to detect structural errors, leakage of administrative text, duplication and unsupported content. c, Retry procedure and auto logging for failed reports, together with the most frequent QC failure flags observed during large-scale processing.

**Primary report parsing.** The system and user prompts below define the role, scope and extraction constraints of the primary parsing agent.

**Primary parsing: system prompt**

```
You are a precise radiology report extraction assistant.  Your task is to segment the
radiology report inside <REPORT>...</REPORT> into EXACTLY 7 sections.

SECTIONS TO EXTRACT (in order):
1.  EXAMINATION
2.  TECHNIQUE
3.  INDICATION
4.  HISTORY
5.  COMPARISON
6.  FINDINGS
7.  IMPRESSION

PARSING RULES:
- Your goal is faithful extraction, NOT rewriting.
- Preserve all clinical meaning exactly.
- Preserve ALL dates exactly as written, regardless of their section.
- You may perform minimal formatting cleanup:
• remove section headers (e.g., "METHODOLOGY:", "ANALYSIS:")
• remove patient/applicant names, institution names, hospital references, provider
references
• remove workflow/admin sentences ("Interpretation will be provided by...", "Results were
sent to...")
• remove duplicated text
• correct spacing, punctuation, and line breaks
• merge fragmented lines into coherent sentences
- DO NOT introduce new clinical findings, interpretations, diagnoses, or details.
- DO NOT change severity, negation, or anatomical descriptors.
- DO NOT hallucinate content not present in the report.
- NEVER delete dates unless they belong to a removed admin/workflow sentence.

SECTION MAPPING LOGIC:

EXAMINATION:
- Exam descriptors such as "MRI BRAIN", "INTRACRANIAL VENOUS ANGIORESONANCE" must be placed
into EXAMINATION.

TECHNIQUE:
- "TECHNIQUE", "METHODOLOGY", "METHOD", "PROTOCOL".

INDICATION:
- "CLINICAL INDICATION", "REASON FOR EXAM", "To evaluate for...", "Rule out...", "follow-up
exam for...", "post-surgical status for...".
- If CLINICAL DATA contains a clear exam purpose ("evaluate for...", "R/O...", "follow-up",
"post-surgical status for..."), map that portion to INDICATION.

HISTORY:
- "CLINICAL DATA", "CLINICAL HISTORY", "HISTORY", and patient symptoms/conditions.
- By default, map "CLINICAL DATA" to HISTORY unless it expresses a reason for exam.
- If INDICATION content include demographic and/or clinical information, move those to
HISTORY.
- Do NOT move text from a labeled HISTORY section into INDICATION.
- HISTORY may contain follow-up statements, symptoms, diagnoses, past procedures, and
relevant clinical context.

COMPARISON:
- Prior exam dates only.  Remove "None" statements.
- If no priors, output "".

FINDINGS vs IMPRESSION DETERMINATION:
- RAW OBSERVATIONS → FINDINGS: Sequence-by-sequence details, measurements, descriptive
text.
```

```
- DIAGNOSTIC SYNTHESIS → IMPRESSION: Final conclusions, recommendations, differential
diagnoses.
- When ambiguous:  If text contains "likely represents," "consistent with," "recommend," or
impression-like statements → IMPRESSION.
- If text describes anatomy without concluding diagnosis → FINDINGS.

EMPTY SECTIONS:
- If a section does not exist → output "".
- If section explicitly states "None", "N/A", or "No prior imaging", output "".

FORMAT RULES:
- Output STRICT JSON with EXACT keys.
- No text outside JSON.
```

**Primary parsing: user prompt**

```
Segment the following radiology report inside <REPORT>...</REPORT> into the required
sections.  Follow the system instructions precisely.
Return ONLY JSON.

<REPORT> {report_text} </REPORT>
```

**Quality-control-guided repair.** To address parsing failures and structural inconsistencies, a secondary prompting stage was used to repair malformed outputs. The model was instructed to correct formatting errors and restore missing or improperly segmented sections without introducing new clinical content. This step ensured consistency with the predefined schema while maintaining fidelity to the original report.

**Repair pass: system prompt**

```
You are an expert radiology report correction assistant.  Your task is to FIX a previously
parsed radiology report so that it completely satisfies all extraction rules AND all QC
criteria.

============================================================
1.  GOAL OF THIS RETRY PASS
============================================================
You are given:
- The ORIGINAL RAW REPORT
- The CURRENT (INCORRECT) PARSED SECTIONS
- A LIST OF QC FLAGS indicating what is wrong

Your job is to repair ONLY the incorrect portions, generating a corrected version of all
seven sections:

EXAMINATION
TECHNIQUE
INDICATION
HISTORY
COMPARISON
FINDINGS
IMPRESSION

============================================================
2.  EXTRACTION RULES (MUST FOLLOW)
============================================================
- Preserve clinical meaning EXACTLY.
- Preserve ALL dates exactly as written.
- Do NOT add new clinical content.
- Do NOT remove true clinical content.
- You may perform minimal formatting cleanup only:
```

```
* Correct spacing and punctuation
* Merge broken lines
* Remove duplicated text
* Remove headers such as \ANALYSIS:" or \METHODOLOGY:"
* Remove admin/workflow statements
- Every section MUST be a single string value.
- Do NOT output lists or arrays.  NEVER output ["sentence1", "sentence2"].

============================================================
3.  QC-AWARE REPAIR RULES (CRITICAL)
============================================================

A. HANDLING DUPLICATED OR CONFUSED HISTORY vs INDICATION
------------------------------------------------------------
If HISTORY and INDICATION contain the SAME text:
- Determine whether the content describes a reason for exam or true clinical history.
- Place the reason-for-exam into INDICATION.
- Place patient symptoms/history/diagnoses into HISTORY.
- If one section is truly redundant, keep the better-fitting one and empty the other.

If HISTORY contains both history AND exam indication:
- Split correctly into INDICATION and HISTORY.

If INDICATION contains clinical history:
- Move the history portion to HISTORY.

B. DUPLICATED FINDINGS AND IMPRESSION
------------------------------------------------------------
If IMPRESSION repeats FINDINGS:
- Keep the FINDINGS section as-is.
- IMPRESSION should contain ONLY diagnostic synthesis, final conclusions, or clinical
interpretation.
- Remove from IMPRESSION any sentences that are direct restatements of FINDINGS.
- If IMPRESSION contains purely repeated image descriptions with no synthesis, reduce it to
the minimal correct summary.
- If IMPRESSION contains a meaningful list but includes duplicated items, keep the list
structure and only remove clear duplicates.

C. UNEXPECTED OR MISSING SECTIONS
------------------------------------------------------------
- If a section should exist but is empty → fill it using the correct content from the RAW
REPORT.
- If a section should NOT exist but contains text → remove the incorrect content.

D. HEADER LEAKAGE
------------------------------------------------------------
Remove any leaked headers such as:  ANALYSIS:
TECHNIQUE:
FINDINGS:
IMPRESSION:
HISTORY:

============================================================
4.  OUTPUT FORMAT RULES
============================================================
- Output STRICT JSON.
- EXACT keys only:
{
"EXAMINATION":  "...",
"TECHNIQUE":  "...",
"INDICATION":  "...",
"HISTORY":  "...",
"COMPARISON":  "...",
"FINDINGS":  "...",
"IMPRESSION":  "..."
}
```

```
- No additional fields.
- Each value must be a string, not list or object.
- No commentary outside JSON.
```

**Repair pass: user prompt**

```
Below is a radiology report, the previously parsed sections, and the QC flags that describe
what is incorrect.

Use the system instructions to FIX the parsed sections.
Return ONLY corrected JSON.

============================================================
ORIGINAL REPORT
============================================================
{raw_report}

============================================================
CURRENT PARSED OUTPUT
============================================================
{parsed_json}

============================================================
QC FLAGS
============================================================
{qc_flags}

============================================================
YOUR TASK
============================================================
Produce corrected JSON that fully satisfies all rules and resolves all QC issues.
```

**Repair pass sampling parameter schedule.** A scheduled sampling policy was used during retry passes to reduce repeated failure modes by gradually loosening decoding constraints.

**Repair pass: sampling parameter schedule**

```
RETRY_SAMPLING = [
  dict(temperature=0.1, top_p=1.0),
  dict(temperature=0.2, top_p=0.95),
  dict(temperature=0.3, top_p=0.95),
  dict(temperature=0.4, top_p=0.9),
]
```

**Post-repair confidence estimation.** Following parsing and repair, a separate prompt was used to estimate the structural reliability of the processed report. The model was instructed to evaluate whether all required sections were correctly identified and formatted, providing a lightweight confidence signal used for downstream filtering and quality assurance.

**Confidence scoring: system prompt**

```
Return ONLY JSON with a 'confidence' field.
```

#### Confidence scoring: user prompt

```
You are evaluating the correctness of a radiology report sectioning task.

You will be given:
1.  The raw radiology report.
2.  The corrected parsed 7-section JSON.

Evaluate:
- Does each section match the meaning and content of the raw report?
- Is any content hallucinated or missing?
- Are dates preserved?
- Did minimal cleanup rules appear to be followed?

Give a single confidence score from 0.0 to 1.0.
- 1.0 = perfect faithful extraction
- 0.0 = major hallucination or mismatch

Output strictly in JSON:
{{
  "confidence":  <float>
}}

===========================
RAW REPORT:
---------------------------
{raw_report}
---------------------------

CORRECTED PARSED OUTPUT:
---------------------------
{corrected_str}
---------------------------
```

### S2 Prompt design for instruction-tuning dataset.

This section outlines the structured prompts used to construct the instruction-tuning dataset, including the system prompts for both the neuroradiologist and reviewer, as well as the iterative revision process. The prompts were designed to ensure that all generated question-answer (QA) pairs are clinically accurate, evidence-based, and strictly based on the source radiology reports.

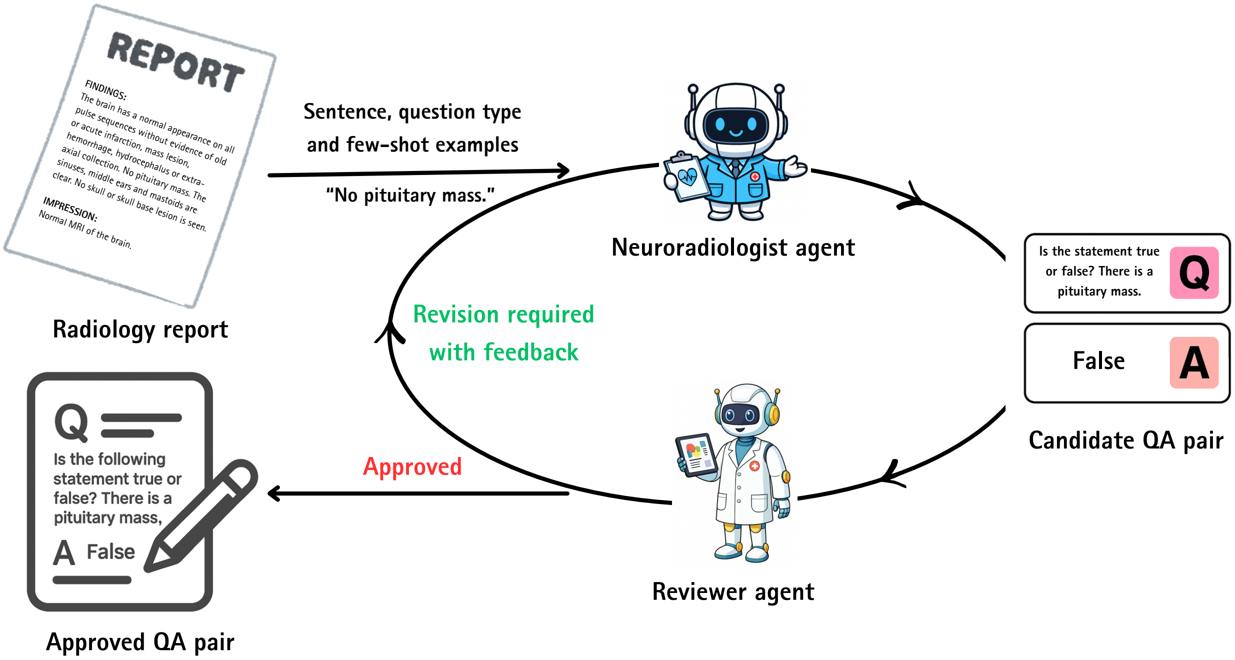


**Figure S2: Overview of the multi-agent system (MAS) for instruction tuning dataset curation.** A neuroradiologist agent generates candidate question–answer (QA) pairs from individual radiology report sentences, conditioned on question type and few-shot exemplars. A second neuroradiologist reviewer agent performs quality control by validating factual consistency and clinical correctness. Drafted pairs requiring correction are iteratively revised using structured feedback until approval, yielding a high-fidelity instruction-tuning corpus.

**Neuroradiologist agent.** This agent was responsible for generating a single clinically grounded QA pair from each input finding sentence and, when needed, revising the pair in response to reviewer feedback.

System prompt: neuroradiologist

```
{ "role":  "system",
"content":  (
  "You are a board-certified neuroradiologist.  Your expertise is in diagnosing and
characterizing abnormalities of the brain, and broadly, disorders of the nervous system
using neuroimaging techniques such as MRI. You must provide precise, clinically accurate,
and evidence-based information."
)
}
```

**User prompt: neuroradiologist**

```
{ "role": "user",
"content": (
  f"Relevant neuroimaging finding: <FINDING>sen</FINDING>\n\n"
  "Your task: Generate ONE question-answer (QA) pair directly based on this neuroimaging
finding.\n\n"
  "Requirements:\n"
  "1. The question must be clinically meaningful and explicitly framed in relation to the
patient's imaging data (e.g., T1, T2, FLAIR, DWI, etc.).\n"
  "2. The answer should be concise, accurate, and derived from the provided finding.\n"
  f"3. The question type should be qtype and here are some examples: format_examples.\n"
  "4. Do NOT introduce new details, speculate or add context that is not present in the
finding.\n\n"
  "Output format:\n"
  "<QUESTION>Your question here</QUESTION>\n"
  "<ANSWER>Your answer here</ANSWER>"
)
}
```

**User prompt: neuroradiologist revision**

```
{ "role": "user",
"content": (
  f"Relevant neuroimaging finding: <FINDING>{sen}</FINDING>\n\n"
  "Previously generated question-answer pair: "
  f"<QUESTION>{question}</QUESTION>\n"
  f"<ANSWER>{answer}</ANSWER>\n"
  f"The feedback from the reviewer is: {feedback}\n\n"
  "Your task: Generate ONE question-answer (QA) pair directly based on this neuroimaging
finding.\n\n"
  "Requirements:\n"
  "1. The question must be clinically meaningful and explicitly framed in relation to the
neuroimaging data (i.e., T1, T2, FLAIR, DWI, etc.).\n"
  "2. The answer should be concise, accurate, and derived from the provided finding.\n"
  f"3. The question type should be {qtype} and here are some examples:
{format_examples}.\n"
  "4. Do NOT introduce new details, speculate or add context that is not present in the
finding.\n\n"
  "Output format:\n"
  "<QUESTION>Your question here</QUESTION>\n"
  "<ANSWER>Your answer here</ANSWER>"
)
}
```

**Reviewer agent.** This agent evaluated whether each generated QA pair was fully supported by the source finding and returned either approval or structured feedback for revision.

**System prompt: reviewer**

```
{ "role": "system",
"content": (
  "You are a board-certified neuroradiologist tasked with rigorously reviewing a single
question-answer (QA) pair produced by another neuroradiologist. Be strict, evidence-based,
and clinically precise. Do NOT introduce any new clinical facts, do NOT rewrite or
generate new QAs, and do NOT infer beyond the provided finding."
)
}
```

**User prompt: reviewer**

```
{
  "role": "user",
  "content": (
    f"Relevant neuroimaging finding: <FINDING>{sen}</FINDING>\n\n"
    "Question-answer pair generated by the neuroradiologist:\n"
    f"<QUESTION>{question}</QUESTION>\n"
    f"<ANSWER>{answer}</ANSWER>\n\n"
    "Please carefully evaluate the question and answer. "
    "If the question-answer pair is fully supported by the provided finding, "
    "respond EXACTLY as:\n<DECISION>APPROVED</DECISION>\n\n"
    "If any problem exists, respond EXACTLY as:\n"
    "<DECISION>REVISION REQUIRED</DECISION>\n"
    "<FEEDBACK>Your feedback here</FEEDBACK>\n"
    "Rules for your evaluation and output:\n"
    "1. Do NOT invent or add new information.\n"
    "2. Do NOT propose or generate new QAs.\n"
    "3. Check that the question is directly answerable from the provided finding.\n"
    "4. Check that the answer is strictly supported (i.e., no hallucination, no
speculation).\n"
    "5. Use only the required tags with no extra text outside them."
  )
}
```

**Few-shot examples for each QA type.** To encourage consistent formatting and high-quality outputs, three few-shot examples were included for each predefined QA type. These examples illustrate the expected question style, answer format, and grounding in the source sentence.

**Few-shot examples for each QA type**

```
1: {
  "Question type": "Factual QA",
  "examples": [
    {
      "sentence": "No mass or acute hemorrhage is detected.",
      "question": "Are there any masses or signs of acute hemorrhage?",
      "answer": "No mass or acute hemorrhage is detected."
    },
    {
      "sentence": "Mastoids are clear.",
      "question": "What is the condition of the mastoid air cells?",
      "answer": "They are clear."
    },
    {
      "sentence": "The vertebral arteries are codominant and unremarkable.",
      "question": "Which characteristics are noted for the vertebral arteries?",
      "answer": "They are codominant and unremarkable."
    }
  ]
},

2: {
  "Question type": "Yes/No QA",
  "examples": [
    {
      "sentence": "Bilateral hippocampal atrophy.",
      "question": "Is there atrophy of the bilateral hippocampi?",
      "answer": "Yes"
    },
    {
      "sentence": "No foci of restricted diffusion to suggest acute infarction.",
      "question": "Is there evidence of acute infarction?",
```

```
      "answer":  "No"
    },
    {
      "sentence":  "Extracranial internal carotid arteries are normal in caliber.",
      "question":  "Do the extracranial internal carotid arteries have a normal caliber?",
      "answer":  "Yes"
    }
  ]
},

3:  {
  "Question type":  "Multiple Choice QA",
  "examples":  [
    {
      "sentence":  "Mastoids are clear.",
      "question":  "Which description applies to the mastoid air cells?  (A) Clear (B)
Opacified",
      "answer":  "A"
    },
    {
      "sentence":  "Major flow voids are unremarkable.",
      "question":  "How are the major flow voids characterized?  (A) Enlarged (B)
Unremarkable (C) Narrowed (D) Unidentified",
      "answer":  "B"
    },
    {
      "sentence":  "Mild mucosal thickening in the ethmoidal air cells.",
      "question":  "What is observed in the ethmoidal air cells?  (A) Clear (B) Mild mucosal
thickening (C) Mass lesion",
      "answer":  "B"
    }
  ]
},

4:  {
  "Question type":  "Classification",
  "examples":  [
    {
      "sentence":  "Mastoids are clear.",
      "question":  "Do the mastoid air cells appear normal or abnormal?",
      "answer":  "Normal"
    },
    {
      "sentence":  "Diffuse generalized parenchymal volume loss with disproportionate volume
loss within the bilateral temporal and parietal lobes.",
      "question":  "How should the parenchymal volume be classified:  preserved or
atrophic?",
      "answer":  "Atrophic"
    },
    {
      "sentence":  "The visualized common carotid arteries and bifurcations are normal
caliber.",
      "question":  "Are the common carotid arteries of normal caliber or abnormal?",
      "answer":  "Normal"
    }
  ]
},

5:  {
  "Question type":  "True/False",
  "examples":  [
    {
      "sentence":  "There is a partially empty sella.",
      "question":  "A partially empty sella is present.  Is this finding True or False?",
      "answer":  "True"
    },
```

```
    {
      "sentence":  "No mass or acute hemorrhage is detected.",
      "question":  "True or False:  There is evidence of acute hemorrhage.",
      "answer":  "False"
    },
    {
      "sentence":  "Bilateral MCAs and ACAs are normal caliber.",
      "question":  "Is the following statement true or false?  Both MCAs and ACAs show
normal caliber.",
      "answer":  "True"
    }
  ]
},

6:  {
  "Question type":  "Extraction",
  "examples":  [
    {
      "sentence":  "The vertebral arteries are codominant and unremarkable.",
      "question":  "How are the vertebral arteries characterized?",
      "answer":  "Codominant and unremarkable."
    },
    {
      "sentence":  "Diffuse generalized parenchymal volume loss with disproportionate volume
loss within the bilateral temporal and parietal lobes.",
      "question":  "What abnormality is observed in the bilateral temporal and parietal
lobes?",
      "answer":  "Disproportionate volume loss."
    },
    {
      "sentence":  "Mild mucosal thickening in the ethmoidal air cells.",
      "question":  "Which finding is described in the ethmoidal air cells?",
      "answer":  "Mild mucosal thickening."
    }
  ]
},

7:  {
  "Question type":  "Opposite Statement (Contradiction)",
  "examples":  [
    {
      "sentence":  "No mass or acute hemorrhage is detected.",
      "question":  "Imaging shows a mass or acute hemorrhage.  Is this statement correct?",
      "answer":  "No"
    },
    {
      "sentence":  "Mastoids are clear.",
      "question":  "The mastoid air cells appear opacified.  Is that true?",
      "answer":  "No"
    },
    {
      "sentence":  "The visualized common carotid arteries and bifurcations are normal
caliber.",
      "question":  "The common carotid arteries are of abnormal caliber.  Does this match
the findings?",
      "answer":  "No"
    }
  ]
},

8:  {
  "Question type":  "Fill-in-the-Blank",
  "examples":  [
    {
      "sentence":  "Mastoids are clear.",
      "question":  "Fill in:  ‘‘Mastoid ___ are clear.’’",
```

```
      "answer":  "Air cells"
    },
    {
      "sentence":  "Diffuse generalized parenchymal volume loss.",
      "question":  "Fill in the missing word:  ‘‘Diffuse generalized parenchymal ___
loss.’’",
      "answer":  "Volume"
    },
    {
      "sentence":  "No focal skull lesion is detected.",
      "question":  "Complete the statement:  ‘‘No ___ skull lesion is detected.’’",
      "answer":  "Focal"
    }
  ]
},

9:  {
  "Question type":  "Cloze (Masked Word)",
  "examples":  [
    {
      "sentence":  "Mastoids are clear.",
      "question":  "The [MASK] of the mastoids are clear.",
      "answer":  "Air cells"
    },
    {
      "sentence":  "There is a partially empty sella.",
      "question":  "There is a partially [MASK] sella.",
      "answer":  "Empty"
    },
    {
      "sentence":  "Bilateral hippocampal atrophy.",
      "question":  "Bilateral hippocampal [MASK].",
      "answer":  "Atrophy"
    }
  ]
},

10:  {
  "Question type":  "Sentence Completion",
  "examples":  [
    {
      "sentence":  "Bilateral hippocampal atrophy.",
      "question":  "Complete the finding:  ‘‘Bilateral ___ ___.’’",
      "answer":  "Hippocampal atrophy"
    },
    {
      "sentence":  "No mass or acute hemorrhage is detected.",
      "question":  "Complete the sentence:  ‘‘No ___ or ___ ___ is detected.’’",
      "answer":  "Mass or acute hemorrhage"
    },
    {
      "sentence":  "There is a partially empty sella.",
      "question":  "Complete:  ‘‘There is a ___ ___ ___.’’",
      "answer":  "Partially empty sella"
    }
  ]
},

11:  {
  "Question type":  "Minimal Pair Discrimination",
  "examples":  [
    {
      "sentence":  "Bilateral hippocampal atrophy.",
      "question":  "Which is correct according to the imaging?  (A) Bilateral hippocampal
atrophy.  (B) Bilateral hippocampal hypertrophy.",
      "answer":  "A"
```

```
    },
    {
      "sentence":  "Mastoids are clear.",
      "question":  "Which matches the sentence?  (A) The mastoid air cells are opacified.
(B) The mastoid air cells are clear.",
      "answer":  "B"
    },
    {
      "sentence":  "No focal skull lesion is detected.",
      "question":  "Which is correct?  (A) No focal skull lesion is detected.  (B) A focal
skull lesion is detected.",
      "answer":  "A"
    }
  ]
},

12:  {
  "Question type":  "Choice Verification",
  "examples":  [
    {
      "sentence":  "Bilateral lens replacements, otherwise the orbits are unremarkable.",
      "question":  "Which option accurately reflects the imaging findings?  (A) Bilateral
lens replacements, otherwise the orbits are unremarkable.  (B) Bilateral lens are intact
with no replacements noted.",
      "answer":  "A"
    },
    {
      "sentence":  "The vertebral arteries are codominant and unremarkable.",
      "question":  "Select the statement that correctly describes the vertebral arteries:
(A) The vertebral arteries are codominant and unremarkable.  (B) The vertebral arteries are
stenotic.",
      "answer":  "A"
    },
    {
      "sentence":  "Diffuse generalized parenchymal volume loss with disproportionate volume
loss within the bilateral temporal and parietal lobes.",
      "question":  "Which choice best matches the reported imaging findings?  (A) Diffuse
generalized parenchymal volume loss with disproportionate loss in bilateral temporal and
parietal lobes.  (B) Normal brain parenchymal volume.",
      "answer":  "A"
    }
  ]
}
```

### S3 Question template for NACC imaging evidence variables.

For each imaging evidence variable derived from the NACC [?] cohort, we defined a fixed set of five semantically equivalent question templates to generate corresponding question-answer (QA) pairs. These templates were designed to probe the same underlying clinical concept using varied phrasing, while preserving identical answer semantics. For each imaging variable, the five templates were paired with categorical labels (Yes, No, Unknown, or no relevant imaging data available), resulting in five QA sets per variable. Across all variables, this yielded 21,975 QA pairs per template. Representative question templates for each imaging evidence variable are listed below.

**NACC imaging evidence question template**

```
{
  'CVDIMAG1':  [
    "Is there evidence of a single strategic infarct on MR imaging?",
```

```
    "Does MR imaging show the presence of a single strategic infarct?",
    "Is a single strategic infarct present in the provided scan(s)?",
    "Are there signs of a single strategic infarct on the scan(s)?",
    "Can a single strategic infarct be identified on the imaging?"
  ],

  'CVDIMAG2':  [
    "Is there evidence of multiple infarcts on MR imaging?",
    "Does the imaging show the presence of multiple infarcts?",
    "Are multiple infarcts present in the provided scan(s)?",
    "Does the provided scan(s) reveal multiple infarcts?",
    "Can multiple infarcts be detected on the imaging?"
  ],

  'HIPPATR': [
    "Is there evidence of hippocampal atrophy on MR imaging?",
    "Does the imaging show hippocampal atrophy?",
    "Is hippocampal atrophy visible on imaging?",
    "Are there signs of hippocampal atrophy on the provided scan(s)?",
    "Does the hippocampus appear atrophied on MRI?"
  ],

  'IMAGLAC': [
    "Are there lacunar infarcts visible on MR imaging?",
    "Does the imaging show evidence of lacunar infarcts?",
    "Are lacunar infarcts present in the imaging?",
    "Does the scan(s) reveal lacunar infarcts?",
    "Is there evidence of small vessel disease with lacunar infarcts?"
  ],

  'IMAGMACH': [
    "Are there macrohemorrhages visible on MR imaging?",
    "Does the imaging show evidence of macrohemorrhages?",
    "Are macrohemorrhages present in the imaging?",
    "Does the scan(s) reveal macrohemorrhages?",
    "Is there evidence of large hemorrhages on MRI?"
  ],

  'IMAGMICH': [
    "Are there microhemorrhages visible on MR imaging?",
    "Does the imaging show evidence of microhemorrhages?",
    "Are microhemorrhages present in the imaging?",
    "Does the scan(s) reveal microhemorrhages?",
    "Is there evidence of small hemorrhages on MRI?"
  ],

  'IMAGEWMH': [
    "Is there extensive white matter hyperintensity on the imaging?",
    "Does the imaging show extensive white matter hyperintensity?",
    "Are there extensive white matter hyperintensities visible?",
    "Does the scan(s) reveal extensive white matter hyperintensity?",
    "Is there evidence of extensive white matter hyperintensities on the imaging?"
  ],

  'INFNETW': [
    "Is there evidence of cystic infarction(s) in cognitive networks on the imaging?",
    "Does the MR imaging show cystic infarction(s) in cognitive networks?",
    "Are there cystic infarction(s) in cognitive networks visible on the imaging?",
    "Does the scan(s) reveal cystic infarction(s) in cognitive networks?",
    "Is there evidence of cystic infarction(s) in cognitive networks on MRI?"
  ],

  'INFWMH': [
    "Is there evidence of combined infarction, extensive WMH, and executive impairment on
the imaging?",
    "Does MRI show combined infarction, extensive WMH, and executive impairment?",
```

```
    "Are there signs of combined infarction, extensive WMH, and executive impairment?",
    "Does the scan(s) reveal combined infarction, extensive WMH, and executive impairment?",
    "Is there evidence of the combined infarction, extensive WMH, and executive impairment
on MRI?"
  ],

  'STKIMAG': [
    "Is stroke confirmed by neuroimaging?",
    "Does the imaging confirm the presence of a stroke?",
    "Is there neuroimaging evidence of stroke?",
    "Does the scan(s) confirm a stroke?",
    "Are there signs of stroke visible on MRI?"
  ],

  'MRFTLD': [
    "Is there structural imaging evidence of frontal or anterior temporal atrophy?",
    "Does the imaging show frontal or anterior temporal atrophy (FTLD)?",
    "Is there evidence of frontal or anterior temporal atrophy on the imaging?",
    "Does the scan(s) reveal frontal or anterior temporal atrophy?",
    "Are there signs of atrophy in the frontal or anterior temporal lobes on MRI?"
  ]
}
```

**Table S1: Keyword term sets for Gradient Health `StudyDescription` screening.** Term sets used to classify visits as intracranial (include) versus non-brain-only (exclude). Matching was performed on lowercased text using regular-expression search; visits were retained if any inclusion term matched, and excluded if no inclusion term matched and any exclusion term matched.

| Field | Inclusion terms (intracranial) | Exclusion terms (non-brain-only) |
|---|---|---|
| `Study-description` | crˆgeral; cran(i/io/eo); crân; crânio/crân; cranium; cerebral; brain; encef; head; hipofis; pituit; sela; turcica; iac; arterial; angio | cervical; spine; coluna; lombo; sacra; lumbar; dorsal; tspine; face; seios; sinus; orbit; mastoid; ouvido; tempora; atm; mandibul; pelv; prostate; leg; hip; whole body; plexus |

**Table S2: Rule hierarchy for MRI series labeling and inclusion in model inputs.** Deterministic rule set used to assign each imaging series to a canonical label using file naming and available sidecar metadata. Metadata text was constructed from the file name and, when available, JSON fields (`SeriesDescription`, `ProtocolName`, `ScanOptions`, `ImageType`); `BidsGuess` and `EchoTime` were used by specialized rules. Rules were applied sequentially (first matching rule applied). Series labeled `LOCALIZER` and `OTHER` were excluded from model inputs as non-diagnostic; `REFORMAT` was retained because for a subset of visits diagnostic content was available only as reconstructions/reformats.

| Step | Rule category | Decision logic and signals (examples) | Output / input use |
|---|---|---|---|
| 1 | Duplicate/equalized exclusion | File-name heuristics (e.g., suffix `_eq_1`). | `OTHER` (excluded) |
| 2 | Diffusion via gradient files | `.bval` → diffusion; `.bvec` with non-trivial gradients → `DTI`. | `DWI`/`DTI` (included) |
| 3 | Localizer detection | Scout/survey/localizer patterns. | `LOCALIZER` (excluded) |
| 4 | Reformat detection | Reformat/recon patterns; MPRAGE exception → `T1`. | `REFORMAT` (included) |
| 5 | Primary diagnostic inference | Ordered pattern matching (see Supplementary Table 2). | Diagnostic label (included) |
| 6 | Field map detection | Field map indicators; TE$<$ 6ms + FFE signature. | `FIELDMAP` (excluded) |
| 7 | Pituitary DCE detection | Dynamic/pituitary/sellar patterns. | `DCE_PITUITARY` (included) |
| 8 | MRV refinement | Venography patterns and vendor hints. | `MRV` (included) |
| 9 | ScanOptions fallback | `ScanOptions` refine `T1` vs `T1CE`. | `T1`/`T1CE` (included) |
| 10 | Fallback | No rule matched. | `UNKNOWN_DIAGNOSTIC` (excluded) |

**Table S3: Frequent modality hallucination templates identified by the detection framework for the baseline Q8B model under greedy decoding on the BSH cohort.** Templates are grouped by hallucination setting, modality family, polarity and template subtype. Templates are normalized by abstracting variable elements (for example, laterality and numeric values). Counts indicate the number of flagged occurrences.

| Family | Polarity | Template (normalized) | Count |
|---|---|---|---|
| | | *Natural-missing hallucinations* | |
| POST_CONTRAST_T1 | NEG | no focal lesions, edema, or enhancement are identified | 11 |
| POST_CONTRAST_T1 | NEG | no abnormal enhancement is seen | 7 |
| POST_CONTRAST_T1 | NEG | no abnormal enhancement is present due to the absence of contrast administration | 5 |
| POST_CONTRAST_T1 | NEG | no focal lesions, edema, or abnormal enhancement are identified | 4 |
| POST_CONTRAST_T1 | NEG | no abnormal enhancement is present | 4 |
| POST_CONTRAST_T1 | NEG | no enhancement is present due to the absence of contrast administration | 4 |
| POST_CONTRAST_T1 | NEG | no new enhancing lesions are identified | 4 |
| SUSCEPTIBILITY | NEG | there is no evidence of hemorrhage on susceptibility-weighted imaging | 9 |
| VASCULAR | NEG | mra reveals no significant stenosis or occlusion of major intracranial vessels | 2 |
| POST_CONTRAST_T1 | POS | the largest lesion is located in the <LAT> frontal lobe, measuring approximately <NUM> <UNIT> in maximum diameter, with heterogeneous enhancement and surrounding vasogenic edema | 6 |
| POST_CONTRAST_T1 | POS | the brain mri demonstrates multiple enhancing lesions consistent with metastatic disease | 5 |
| | | *Counterfactual hallucinations* | |
| DIFFUSION | NEG | diffusion-weighted imaging shows no restricted diffusion | 27 |
| DIFFUSION | NEG | no focal lesions, infarcts, or areas of restricted diffusion are identified | 25 |
| DIFFUSION | NEG | there is no abnormal enhancement, edema, or restricted diffusion | 8 |
| DIFFUSION | NEG | no focal lesions, edema, or restricted diffusion are identified | 6 |
| VASCULAR | NEG | mra reveals no significant stenosis or occlusion of major intracranial vessels | 7 |
| POST_CONTRAST_T1 | NEG | no abnormal enhancement is seen following contrast administration | 10 |
| POST_CONTRAST_T1 | NEG | no enhancement is seen following contrast administration | 4 |
| POST_CONTRAST_T1 | NEG | there is no abnormal enhancement following contrast administration | 4 |
| POST_CONTRAST_T1 | NEG | there is no abnormal enhancement on post-contrast sequences | 4 |
| POST_CONTRAST_T1 | POS | the largest lesion is located in the <LAT> frontal lobe, measuring approximately <NUM> <UNIT> in maximum diameter, with heterogeneous enhancement and surrounding vasogenic edema | 11 |

*Continued on next page*

| **Family** | **Polarity** | **Template (normalized)** | **Count** |
|---|---|---|---|
| POST_CONTRAST_T1 | POS | the brain mri demonstrates multiple enhancing lesions consistent with metastatic disease | 7 |
| DIFFUSION | POS | the lesion demonstrates restricted diffusion on dwi sequences and corresponding hyperintensity on adc maps | 3 |
| | | *Illustrative vascular and susceptibility examples* | |
| VASCULAR | POS | occlusion of the proximal <LAT> middle cerebral arteriole confirmed by mra | 2 |
| VASCULAR | POS | normal vascular anatomy on mra | 2 |
| SUSCEPTIBILITY | NEG | there is no evidence of hemorrhage on susceptibility-weighted imaging | 4 |
| SUSCEPTIBILITY | NEG | no abnormal perivascular spaces or microbleeds are seen | 2 |
| SUSCEPTIBILITY | POS | there is evidence of hemosiderin deposition in the subcortical white matter bilaterally, particularly in the frontal lobes | 1 |

**Table S4: Performance on multiple-choice visual question-answering tasks across cohorts.** Accuracy, precision, and recall for each model variant (Q8B baseline, QA instruction-tuned (IT), and RRG supervised fine-tuned (SFT)) evaluated on multiple-choice question types across four cohorts (GH, BMC, LHMC, and BSH). Metrics are reported per cohort and averaged with standard deviation at the bottom. The best result for each metric is shown in bold.

| Model | **GH** ($n = 39,340$) | | **BMC** ($n = 27,156$) | | **LHMC** ($n = 17,327$) | | **BSH** ($n = 3,462$) | |
|---|---|---|---|---|---|---|---|---|
| | Precision | Recall | Precision | Recall | Precision | Recall | Precision | Recall |
| **Choice verification** | | | | | | | | |
| Q8B | 0.834 | 0.827 | 0.854 | 0.848 | 0.860 | 0.854 | 0.787 | 0.778 |
| Q8B + IT | 0.849 | 0.786 | 0.883 | 0.848 | 0.866 | 0.819 | 0.823 | 0.734 |
| Q8B + IT + SFT | 0.925 | 0.916 | 0.938 | 0.932 | 0.915 | 0.908 | 0.914 | 0.904 |
| **Minimal pair discrimination** | | | | | | | | |
| Q8B | 0.823 | 0.797 | 0.836 | 0.823 | 0.839 | 0.815 | 0.804 | 0.766 |
| Q8B + IT | 0.964 | 0.962 | 0.952 | 0.947 | 0.927 | 0.926 | 0.927 | 0.918 |
| Q8B + IT + SFT | 0.956 | 0.955 | 0.950 | 0.947 | 0.923 | 0.923 | 0.929 | 0.925 |
| **Multiple choice QA** | | | | | | | | |
| Q8B | 0.815 | 0.779 | 0.833 | 0.805 | 0.799 | 0.767 | 0.850 | 0.835 |
| Q8B + IT | 0.951 | 0.948 | 0.939 | 0.936 | 0.927 | 0.921 | 0.980 | 0.979 |
| Q8B + IT + SFT | 0.942 | 0.939 | 0.944 | 0.941 | 0.935 | 0.931 | 0.957 | 0.957 |
| **Opposite statement (contradiction)** | | | | | | | | |
| Q8B | 0.533 | 0.523 | 0.589 | 0.570 | 0.631 | 0.603 | 0.451 | 0.458 |
| Q8B + IT | 0.951 | 0.920 | 0.945 | 0.925 | 0.921 | 0.901 | 0.942 | 0.937 |
| Q8B + IT + SFT | 0.927 | 0.927 | 0.930 | 0.928 | 0.907 | 0.906 | 0.916 | 0.915 |
| **True/False** | | | | | | | | |
| Q8B | 0.733 | 0.705 | 0.752 | 0.730 | 0.768 | 0.727 | 0.799 | 0.792 |
| Q8B + IT | 0.971 | 0.922 | 0.973 | 0.928 | 0.962 | 0.914 | 0.982 | 0.898 |
| Q8B + IT + SFT | 0.956 | 0.932 | 0.956 | 0.940 | 0.936 | 0.909 | 0.970 | 0.937 |
| **Yes/No QA** | | | | | | | | |
| Q8B | 0.697 | 0.659 | 0.693 | 0.673 | 0.718 | 0.687 | 0.699 | 0.696 |
| Q8B + IT | 0.979 | 0.974 | 0.982 | 0.978 | 0.953 | 0.946 | 0.965 | 0.962 |
| Q8B + IT + SFT | 0.977 | 0.976 | 0.975 | 0.975 | 0.949 | 0.948 | 0.995 | 0.995 |
| **Average** (± std) | | | | | | | | |
| Q8B | 0.739 ± 0.115 | 0.715 ± 0.113 | 0.759 ± 0.104 | 0.742 ± 0.106 | 0.769 ± 0.085 | 0.742 ± 0.091 | 0.732 ± 0.146 | 0.721 ± 0.136 |
| Q8B + IT | 0.944 ± 0.048 | 0.919 ± 0.068 | 0.946 ± 0.035 | 0.927 ± 0.043 | 0.926 ± 0.034 | 0.904 ± 0.044 | 0.937 ± 0.060 | 0.905 ± 0.089 |
| Q8B + IT + SFT | **0.947** ± **0.020** | **0.941** ± **0.022** | **0.949** ± **0.016** | **0.944** ± **0.017** | **0.927** ± **0.015** | **0.921** ± **0.017** | **0.947** ± **0.033** | **0.939** ± **0.033** |

**Table S5: Performance on free-form visual question-answering tasks across cohorts.** BLEU-4, ROUGE-L, METEOR, and BERT-F1 for each model variant (Q8B baseline, QA instruction-tuned (IT), and RRG supervised fine-tuned (SFT)) evaluated on free-form question types across four cohorts (GH, BMC, LHMC, and BSH). Metrics are reported per cohort and averaged with standard deviation at the bottom. The best result for each metric is shown in bold.

| **Model** | **GH** ($n = 52,004$) | | | **BMC** ($n = 26,455$) | | | **LHMC** ($n = 15,074$) | | | **BSH** ($n = 3,536$) | | |
|---|---|---|---|---|---|---|---|---|---|---|---|---|
| | BLEU-4 | ROUGE-L | METEOR | BLEU-4 | ROUGE-L | METEOR | BLEU-4 | ROUGE-L | METEOR | BLEU-4 | ROUGE-L | METEOR |
| **Classification** | | | | | | | | | | | | |
| Q8B | 0.087 | 0.464 | 0.242 | 0.087 | 0.460 | 0.242 | 0.051 | 0.274 | 0.147 | 0.132 | 0.720 | 0.367 |
| Q8B + IT | 0.166 | 0.864 | 0.458 | 0.150 | 0.775 | 0.414 | 0.103 | 0.538 | 0.290 | 0.163 | 0.893 | 0.458 |
| Q8B + IT + SFT | 0.165 | 0.857 | 0.455 | 0.153 | 0.776 | 0.421 | 0.100 | 0.533 | 0.282 | 0.163 | 0.894 | 0.460 |
| **Cloze (masked word)** | | | | | | | | | | | | |
| Q8B | 0.042 | 0.220 | 0.131 | 0.046 | 0.219 | 0.137 | 0.048 | 0.248 | 0.141 | 0.049 | 0.266 | 0.146 |
| Q8B + IT | 0.111 | 0.542 | 0.315 | 0.080 | 0.415 | 0.235 | 0.083 | 0.423 | 0.249 | 0.090 | 0.507 | 0.256 |
| Q8B + IT + SFT | 0.102 | 0.490 | 0.288 | 0.074 | 0.376 | 0.217 | 0.079 | 0.392 | 0.238 | 0.057 | 0.325 | 0.166 |
| **Extraction** | | | | | | | | | | | | |
| Q8B | 0.036 | 0.208 | 0.228 | 0.040 | 0.189 | 0.215 | 0.029 | 0.197 | 0.204 | 0.032 | 0.229 | 0.261 |
| Q8B + IT | 0.045 | 0.289 | 0.179 | 0.025 | 0.212 | 0.138 | 0.027 | 0.213 | 0.132 | 0.040 | 0.339 | 0.185 |
| Q8B + IT + SFT | 0.040 | 0.304 | 0.178 | 0.028 | 0.264 | 0.168 | 0.025 | 0.234 | 0.137 | 0.036 | 0.353 | 0.183 |
| **Factual QA** | | | | | | | | | | | | |
| Q8B | 0.062 | 0.332 | 0.416 | 0.077 | 0.342 | 0.425 | 0.059 | 0.316 | 0.400 | 0.047 | 0.315 | 0.394 |
| Q8B + IT | 0.107 | 0.352 | 0.326 | 0.058 | 0.256 | 0.227 | 0.054 | 0.259 | 0.223 | 0.138 | 0.388 | 0.367 |
| Q8B + IT + SFT | 0.130 | 0.422 | 0.421 | 0.100 | 0.352 | 0.340 | 0.075 | 0.333 | 0.329 | 0.157 | 0.440 | 0.452 |
| **Fill-in-the-blank** | | | | | | | | | | | | |
| Q8B | 0.059 | 0.314 | 0.178 | 0.063 | 0.332 | 0.193 | 0.071 | 0.373 | 0.226 | 0.047 | 0.255 | 0.142 |
| Q8B + IT | 0.122 | 0.629 | 0.341 | 0.103 | 0.549 | 0.300 | 0.098 | 0.524 | 0.290 | 0.075 | 0.420 | 0.222 |
| Q8B + IT + SFT | 0.113 | 0.585 | 0.316 | 0.099 | 0.532 | 0.290 | 0.091 | 0.488 | 0.271 | 0.061 | 0.337 | 0.173 |
| **Sentence completion** | | | | | | | | | | | | |
| Q8B | 0.040 | 0.160 | 0.120 | 0.050 | 0.214 | 0.153 | 0.045 | 0.182 | 0.146 | 0.030 | 0.126 | 0.108 |
| Q8B + IT | 0.133 | 0.412 | 0.307 | 0.104 | 0.361 | 0.271 | 0.096 | 0.341 | 0.262 | 0.067 | 0.345 | 0.201 |
| Q8B + IT + SFT | 0.118 | 0.389 | 0.288 | 0.089 | 0.348 | 0.251 | 0.091 | 0.340 | 0.263 | 0.059 | 0.276 | 0.168 |
| **Average** (± std) | | | | | | | | | | | | |
| Q8B | 0.054 ± 0.019 | 0.283 ± 0.110 | 0.219 ± 0.108 | 0.061 ± 0.019 | 0.292 ± 0.104 | 0.228 ± 0.104 | 0.050 ± 0.014 | 0.265 ± 0.072 | 0.211 ± 0.099 | 0.056 ± 0.038 | 0.318 ± 0.206 | 0.236 ± 0.123 |
| Q8B + IT | **0.114 ± 0.040** | **0.515 ± 0.212** | 0.321 ± 0.089 | 0.087 ± 0.043 | 0.428 ± 0.208 | 0.264 ± 0.092 | 0.077 ± 0.030 | 0.383 ± 0.135 | 0.241 ± 0.059 | **0.095 ± 0.046** | **0.482 ± 0.210** | **0.282 ± 0.108** |
| Q8B + IT + SFT | 0.111 ± 0.041 | 0.508 ± 0.196 | **0.324 ± 0.101** | **0.090 ± 0.041** | **0.441 ± 0.186** | **0.281 ± 0.090** | **0.077 ± 0.027** | **0.387 ± 0.110** | **0.254 ± 0.064** | 0.089 ± 0.056 | 0.438 ± 0.230 | 0.267 ± 0.147 |

**Table S6: Performance on interpreting neuroimaging abnormalities.** Evaluation of the vision-language models on a curated multi-choice visual question-answering (VQA) benchmark derived from the National Alzheimer's Coordinating Center (NACC) cohort. Questions were generated from structured imaging variables and designed to require implicit cross-sequence reasoning without cueing specific modalities. Metrics (accuracy, precision, recall and F1-score) are reported across four abnormality categories (infarct(s), hemorrhage(s), atrophy and white-matter hyperintensity), with standard deviations in parentheses. Results are shown for the baseline Q8B model, after QA instruction tuning (Q8B + IT), and after report-specific supervised fine-tuning (Q8B + IT + SFT). All metrics are higher-is-better; bold values indicate the best performance in each column within each category.

| Abnormality | Model | Accuracy | Precision | Recall | F1-score |
|---|---|---|---|---|---|
| Infarct(s) | Q8B | 0.181 0.048 | 0.121 0.039 | 0.177 0.049 | 0.136 0.024 |
| | Q8B + IT | **0.328** 0.051 | **0.220** 0.030 | **0.320** 0.051 | **0.200** 0.019 |
| | Q8B + IT + SFT | 0.265 0.037 | 0.126 0.025 | 0.296 0.037 | 0.175 0.015 |
| Hemorrhage(s) | Q8B | **0.942** 0.018 | 0.346 0.039 | 0.503 0.018 | 0.337 0.053 |
| | Q8B + IT | 0.920 0.013 | **0.526** 0.060 | 0.534 0.013 | **0.529** 0.071 |
| | Q8B + IT + SFT | 0.883 0.011 | 0.523 0.047 | **0.553** 0.011 | 0.525 0.052 |
| Atrophy | Q8B | 0.675 0.057 | 0.436 0.058 | 0.601 0.057 | 0.414 0.076 |
| | Q8B + IT | **0.811** 0.093 | 0.405 0.500 | 0.500 0.093 | **0.448** 0.127 |
| | Q8B + IT + SFT | 0.804 0.065 | **0.419** 0.062 | **0.542** 0.064 | 0.359 0.087 |
| White matter hyperintensity | Q8B | **0.304** 0.067 | 0.415 0.075 | 0.348 0.069 | **0.236** 0.044 |
| | Q8B + IT | 0.284 0.039 | 0.337 0.034 | **0.423** 0.041 | 0.234 0.016 |
| | Q8B + IT + SFT | 0.081 0.015 | **0.441** 0.061 | 0.359 0.015 | 0.087 0.011 |
